# Peripheral immune biomarkers, brain imaging phenotypes and treatment response in major depressive disorder

**DOI:** 10.64898/2026.11.03.26367072

**Authors:** Benjamin Eggerstorfer, Patrick Altmann, Matej Murgaš, Markus Ponleitner, Godber M Godbersen, Manfred Klöbl, Jakob Unterholzner, Leo R Silberbauer, Chrysoula Vraka, Cecile Philippe, Lukas Nics, Andreas Hahn, Marcus Hacker, Gregor Gryglewski, Rupert Lanzenberger

## Abstract

Immune dysregulation is implicated in major depressive disorder (MDD), but its links to clinical features and *in vivo* brain phenotypes remain uncertain. We examined ten plasma biomarkers in 36 unmedicated MDD patients and 48 control participants using high-sensitivity immune profiling at baseline, alongside PET/MR imaging of brain serotonin transporter (SERT) binding and regional structural measures. Patients underwent three months of antidepressant treatment and were followed up with MR imaging. Glial fibrillary acidic protein (GFAP) was higher in MDD patients (β = 0.66, 95% CI 0.22 - 1.11), and interleukin-33 (IL-33) correlated with subcortical volumes across all participants (β = 0.22, 95% CI 0.07 - 0.38). No marker correlated with SERT binding or cortical thickness after correction, and neither SERT binding nor brain structural measures differed by diagnosis. In the longitudinal analysis, interleukin-13 (IL-13) was associated with greater antidepressant response during first-line treatment with escitalopram (β = 1.09, 95% CI 0.56 - 1.61). IL-15 and IL-13 showed nominal associations with hippocampal volume increase after treatment. These findings support astroglial involvement in MDD and link peripheral immune variation to subcortical brain structure. IL-13 is a candidate prognostic marker of response to selective serotonin reuptake inhibitors. Its tentative association with longitudinal changes in hippocampal volume suggests that immune processes in MDD may be related to both the capacity for clinical improvement and brain structural plasticity during treatment.

## Introduction

Major depressive disorder (MDD) is a clinically and biologically heterogeneous disorder for which current diagnostic categories provide little information about underlying pathophysiology or treatment outcome. This heterogeneity has constrained biomarker discovery and may explain why inflammatory, neuroimaging, and molecular measures frequently distinguish patients from healthy individuals at the group level, yet rarely achieve individual-level specificity (Abi-Dargham et al., 2023). Next to the assessment of group differences in biomarkers of MDD, mapping relationships between immune profiles, clinical, and neurobiological measures may clarify how different pathophysiological processes interlock in depression and lead to the development of more effective stratification methods for targeted interventions.

Substantial evidence implicates immune dysregulation in MDD. Meta-analyses have identified higher circulating C-reactive protein (CRP), interleukin (IL)-6, tumor necrosis factor (TNF), and related inflammatory mediators in depression (Köhler et al., 2018; Osimo et al., 2020). Additionally, translocator protein positron emission tomography (PET) studies show altered neuroimmune activity in corticolimbic regions (De Picker et al., 2023; Eggerstorfer et al., 2022). Longitudinal studies further suggest bidirectional associations between inflammation and depression: higher CRP and IL-6 predict subsequent depressive symptoms, while depression may itself contribute to later increases in these markers (Colasanto et al., 2020; Mac Giollabhui et al., 2021). Immune alterations may therefore contribute to depressive pathophysiology while also being reinforced by the course and consequences of illness.

Immune involvement is unlikely to be uniform across MDD. Inflammatory markers have been associated more consistently with anhedonia, fatigue, psychomotor slowing, sleep disturbance, and appetite change than with overall symptom severity (Kofod et al., 2022; Milaneschi et al., 2021). This has led to proposals for an inflammation-related subtype characterized by elevated inflammatory activity and prominent neurovegetative and reward-related symptoms (Jha et al., 2025; Miller, 2025). Inflammation may also contribute to variation in antidepressant outcomes. Higher baseline inflammatory activity has been associated with poorer response to standard antidepressants (Bufalino et al., 2013; Carvalho et al., 2013). But findings vary across markers and treatments, and changes in inflammatory markers do not necessarily follow symptom improvement (Kofod et al., 2022). Whether peripheral markers provide information about subsequent treatment outcome beyond their association with diagnostic status or baseline symptom severity therefore remains unresolved.

Understanding how circulating markers relate to brain biology is another key challenge. Beyond their inflammatory roles, cytokines can modulate synaptic function, with effects that vary across cellular targets and neural circuits (Salvador et al., 2021; Wheeler and Quintana, 2025). Peripheral immune signals can influence the central nervous system via endothelial and vagal pathways (Huerta et al., 2025; Jin et al., 2024), changes in the blood-brain barrier (Huang et al., 2021; Tyagi and Bartley, 2026), immune-cell trafficking (Li et al., 2018; Thomson et al., 2020), kynurenine metabolism (Paul et al., 2022; Savitz, 2020), oxidative and nitrosative stress (di Penta et al., 2013), and shifts in glial activity (Miller and Raison, 2016; Shen et al., 2026; Upthegrove et al., 2025).

Serotonergic regulation may represent one interface between immune activity and antidepressant response, but direct human evidence linking cytokines to *in vivo* SERT availability remains limited. Cytokines may influence serotonergic signaling through tryptophan-kynurenine metabolism, SERT regulation, and neuroplastic pathways (Savitz, 2020; Zhu et al., 2006), while antidepressant treatment has been associated with changes in immune activity (Bleibel et al., 2025; Köhler et al., 2018; Wang et al., 2019).

By contrast, peripheral inflammation has been more consistently associated with structural variation in cortical and limbic regions, including the hippocampus, which is sensitive to inflammatory and stress-related processes and may undergo structural changes during antidepressant treatment (Cordero et al., 2026; Evans et al., 2024; Gryglewski et al., 2021; Popovic et al., 2026; Schmaal et al., 2016). However, immune markers, SERT availability, brain structure, and treatment-related structural change have rarely been examined together.

In the present study, we combined high-sensitivity profiling of circulating immune and glial markers with prospective assessment of antidepressant response, [^11^C]DASB PET, and structural MRI in a thoroughly characterized case-control cohort. By evaluating immune associations across complementary clinical and neurobiological phenotypes, we aimed to determine whether immune markers show generalized associations across MDD-related phenotypes or more selective relationships with diagnosis, treatment outcome, serotonergic transmission, or brain structure.

## Methods and materials

### Participants and study design

Analyses included 36 patients and 48 healthy controls (Table 1), who were partially used in previous studies by our group (Gryglewski et al., 2019; Klöbl et al., 2025, 2020; Seiger et al., 2021; Silberbauer et al., 2022; Zadka et al., 2025). All participants were non-smokers. MDD patients were free from antidepressant treatment for at least three months before inclusion and had no comorbid psychiatric or somatic illnesses. Patients were recruited at the Department of Psychiatry and Psychotherapy at the Medical University of Vienna (Figure 1A). Diagnosis was confirmed using the Structured Clinical Interview for DSM-IV Axis I disorders (SCID-I). Depression severity was assessed using the Montgomery-Åsberg Depression Rating Scale (MADRS), Hamilton Rating Scale for depression 17-item (HDRS_17_), and Beck Depression Inventory (BDI). Participants underwent baseline [^11^C]DASB PET/MR scans to measure SERT binding potential (BP_P_) and brain structure, as in our previous work (see supplement). Patients then received open-label antidepressant treatment starting with escitalopram 10 mg/day and biweekly follow-up for three months, allowing dose escalation or medication switch in case of non-response or intolerance. Patients underwent an additional MRI scan 12 weeks after treatment initiation. Further treatment details are reported elsewhere (Klöbl et al., 2025, 2020; Seiger et al., 2021; Silberbauer et al., 2022; Zadka et al., 2025). All participants gave written informed consent, and the study was approved by the local ethics committee, conducted according to the Declaration of Helsinki, and registered at ClinicalTrials.gov (NCT02711215).

**Figure 1:**
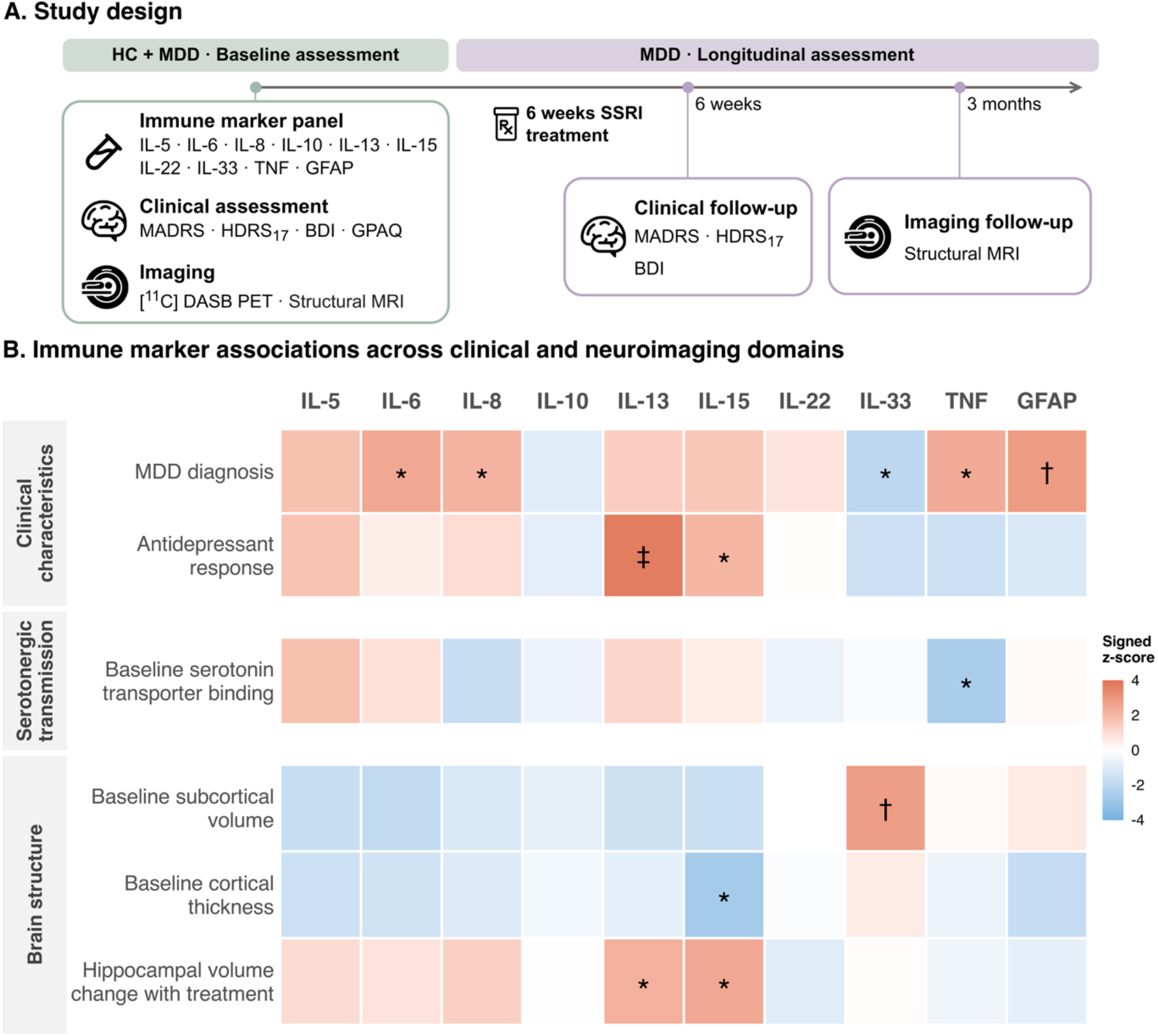
Study design and immune marker associations across clinical and neuroimaging domains. (A) Healthy controls (HC) and patients with major depressive disorder (MDD) underwent baseline clinical assessment, immune marker quantification, [^11^C]DASB positron emission tomography (PET), and structural magnetic resonance imaging (MRI). MDD patients were reassessed clinically and with structural MRI after antidepressant treatment. (B) The heatmap displays signed z-scores for associations across six predefined domains within three categories: clinical characteristics (MDD diagnosis and antidepressant response), serotonergic transmission (baseline SERT binding measured with PET), and brain structure (baseline subcortical brain volumes, cortical thickness, and treatment-related hippocampal volume change measured with MRI). Positive z-scores indicate positive associations or higher levels in MDD, whereas negative z-scores indicate inverse associations or lower levels. For antidepressant response, positive z-scores indicate greater reductions in MADRS scores from baseline to post-treatment. * = nominal *p* < 0.05, † = FDR-adjusted *p* < 0.05 within the corresponding domain, ‡ = FDR-adjusted *p* < .05 across all analyses. BDI, Beck Depression Inventory; FDR, false discovery rate; GFAP, glial fibrillary acidic protein; GPAQ, Global Physical Activity Questionnaire; HDRS_17_, 17-item Hamilton Depression Rating Scale; IL, interleukin; MADRS, Montgomery-Åsberg Depression Rating Scale; SERT, serotonin transporter; SSRI, selective serotonin reuptake inhibitor; TNF, tumor necrosis factor.

**Table 1:** Demographics of the study population. HC, healthy control; MDD, major depressive disorder; BMI, body mass index; CRP, C-reactive protein; GPAQ, Global Physical Activity Questionnaire; MADRS, Montgomery-Åsberg Depression Rating Scale; BDI, Beck Depression Inventory; HDRS_17_, Hamilton Rating Scale for depression 17-item. ^a^ t-test, ^b^ chi-square test, ^c^ datapoint(s) missing

| **Characteristic** | **Overall** | **HC** | **MDD** | **p** |
|---|---|---|---|---|
| N | 84 | 48 | 36 |  |
| Age (years) | 28.2 ± 8.5 | 28.2 ± 8.6 | 28.2 ± 8.4 | 0.99<sup>a</sup> |
| Sex, n (%) |  |  |  | 0.80<sup>b</sup> |
| f | f: 43 (51%) | f: 24 (50%) | f: 19 (53%) |  |
| m | m: 41 (49%) | m: 24 (50%) | m: 17 (47%) |  |
| Weight (kg) | 69.0 ± 12.4 | 69.3 ± 11.5 | 68.6 ± 13.6 | 0.82<sup>a</sup> |
| BMI (kg/m<sup>2</sup>) | 22.8 ± 3.0 | 22.6 ± 2.6 | 23.0 ± 3.5 | 0.53<sup>a</sup> |
| CRP (mg/dL) | 0.1 ± 0.2<sup>c</sup> | 0.1 ± 0.2<sup>c</sup> | 0.1 ± 0.2<sup>c</sup> | 0.77<sup>a</sup> |
| GPAQ total | 20.1 ± 23.0<sup>c</sup> | 23.2 ± 26.3<sup>c</sup> | 14.6 ± 14.5<sup>c</sup> | 0.09<sup>a</sup> |
| MADRS |  |  | 30.6 ± 5.7<sup>c</sup> |  |
| BDI |  |  | 27.7 ± 7.6<sup>c</sup> |  |
| HDRS<sub>17</sub> |  |  | 22.4 ± 4.8 |  |

### Blood sampling and single-molecule array

Venous blood samples were collected during PET/MR scans. Plasma was separated by centrifugation at 3,506 × g for 4 minutes and stored at −80°C until analysis. Samples were transported on dry ice and gently thawed on ice immediately before measurement. Concentrations of inflammatory and neuroglial markers, including interleukins (IL-5, IL-6, IL-8, IL-10, IL-13, IL-15, IL-17A, IL-22, IL-33), TNF, and glial fibrillary acidic protein (GFAP) were measured using single-molecule array technology on the SR-X Analyzer platform (Simoa Quanterix, Lexington, MA, USA) (Rissin et al., 2010). All analyses were performed by an investigator blinded to clinical data, following the manufacturer’s instructions. Samples and calibrators were equilibrated to room temperature, diluted 1:4 in sample diluent, and measured in duplicate in 96-well plates.

Detector reagent and paramagnetic beads were added sequentially to each well, followed by incubation at 30°C and 800rpm for 30 minutes on a Simoa microplate incubator. After automated washing with the Simoa microplate washer, streptavidin β-galactosidase was added, and plates were incubated again at 30°C and 800rpm for 10 minutes. A final wash was followed by drying the plates for 10 minutes before transferring them to the Quanterix SR-X Analyzer for readout. All assay kits and consumables used are detailed in the supplement and were from the same lot where applicable. To reduce inter-assay variability, all samples from a single participant were processed on the same plate. Duplicate measurements underwent quality control as described in the supplementary methods. IL-17A was designated as an exploratory binary detectable marker because its detection rate was below 80% during replicate quality control.

### Statistical analysis

Analyses were performed in R 4.5.3. Continuous variables were inspected using histograms and summary statistics, and skewed variables were log-transformed where appropriate. Immune markers and imaging measures were standardized before analysis. Continuous covariates were mean-centered within the corresponding analysis dataset.

Associations of plasma marker levels were evaluated across six prespecified analysis domains spanning three broader outcome categories: A) Clinical characteristics (MDD diagnosis, antidepressant response); B) Brain serotonergic transmission (baseline PET serotonin transporter binding potential); and C) Brain structure (baseline subcortical volume, baseline cortical thickness, and longitudinal hippocampal change with treatment). Models were fitted separately for each immune marker using marker-specific complete cases. Linear mixed-effects models with participant-specific random intercepts were used for analysis of repeated (regional or longitudinal) measurements.

A) Associations with MDD diagnosis were evaluated using linear regression with immune marker as the outcome and diagnostic group as the predictor. Antidepressant response was assessed using linear mixed-effects models of MADRS scores measured at baseline and after six weeks of treatment. Models included timepoint, immune marker level, and their interaction. Exploratory analyses applied the same approach to BDI and HDRS_17_.

B) Associations of plasma markers with baseline SERT BP_P_ were examined across six predefined PET regions with high SERT availability: anterior cingulate cortex, amygdala, hippocampus, striatum, thalamus, and midbrain. Linear mixed-effects models included immune marker concentration, region (repeated), and diagnostic group, and estimated the average marker association across regions in SERT BP_P_.

C) Baseline structural MRI was analyzed for subcortical volume and cortical thickness.

Subcortical volume analyses included the caudate, amygdala, hippocampus, and nucleus accumbens, based on previous evidence implicating frontostriatal and limbic structural alterations in MDD (Brandl et al., 2022; Serrarens et al., 2025). Left- and right-hemisphere volumes were summed for bilaterally defined regions. Cortical analyses included the rostral and caudal anterior cingulate, insula, and medial orbitofrontal cortex, based on large-scale evidence of depression-related alterations in cingulate, insular, and orbitofrontal thickness (Opel et al., 2019; Schmaal et al., 2017). For each cortical region, bilateral thickness was calculated as the surface area weighted mean of left- and right-hemisphere estimates. Subcortical volumes and cortical thickness measures were standardized for each region, and mixed-effects models for both structural domains included immune marker concentration, region, and diagnostic group, with estimated total intracranial volume (eTIV) included in the subcortical models. Exploratory region × marker interaction models tested for regional heterogeneity. Longitudinal hippocampal analyses were available for MDD patients. Bilateral hippocampal volumes were analyzed in their original units (mm^3^). Primary models included timepoint, immune marker concentration, their interaction, and eTIV, retaining participants with at least one hippocampal measurement. Sensitivity models included complete-pair models. Exploratory Spearman rank correlation assessed the association between six-week MADRS reduction and hippocampal-volume change.

For all domains, the effect of potentially influential covariates was assessed by adjusting for age, sex, BMI, and Global Physical Activity Questionnaire (GPAQ) total score. Benjamini-Hochberg correction was applied within each domain and globally across all 60 marker-domain tests. For cross-domain visualization, two-sided p-values were converted to signed z-scores according to effect direction (Figure 1B).

Exploratory analyses examined routine blood parameters across domains and their correlations with immune markers, adjusted for diagnostic group (Supplementary Figure S4). Parameters included CRP, fibrinogen, total leukocytes, and absolute neutrophil, eosinophil, lymphocyte, monocyte, and platelet counts.

## Results

### Clinical characteristics

The case-control immune profiling cohort comprised 84 participants, including 36 patients with major depressive disorder (MDD) and 48 healthy controls (Table 1). Group differences in age, sex, and BMI were negligible. Due to modality-specific data availability, sample sizes varied across analyses (Supplementary Table S3).

GFAP concentrations were higher in MDD patients than in healthy controls (β = 0.66, 95% CI 0.22 to 1.10, *p*_FDR,domain_ = 0.04, Figure 2A), although the association did not survive global correction (*p*_FDR,global_ = 0.1). TNF, IL-6, and IL-8 were nominally higher in MDD, whereas IL-33 was nominally lower.

**Figure 2.**
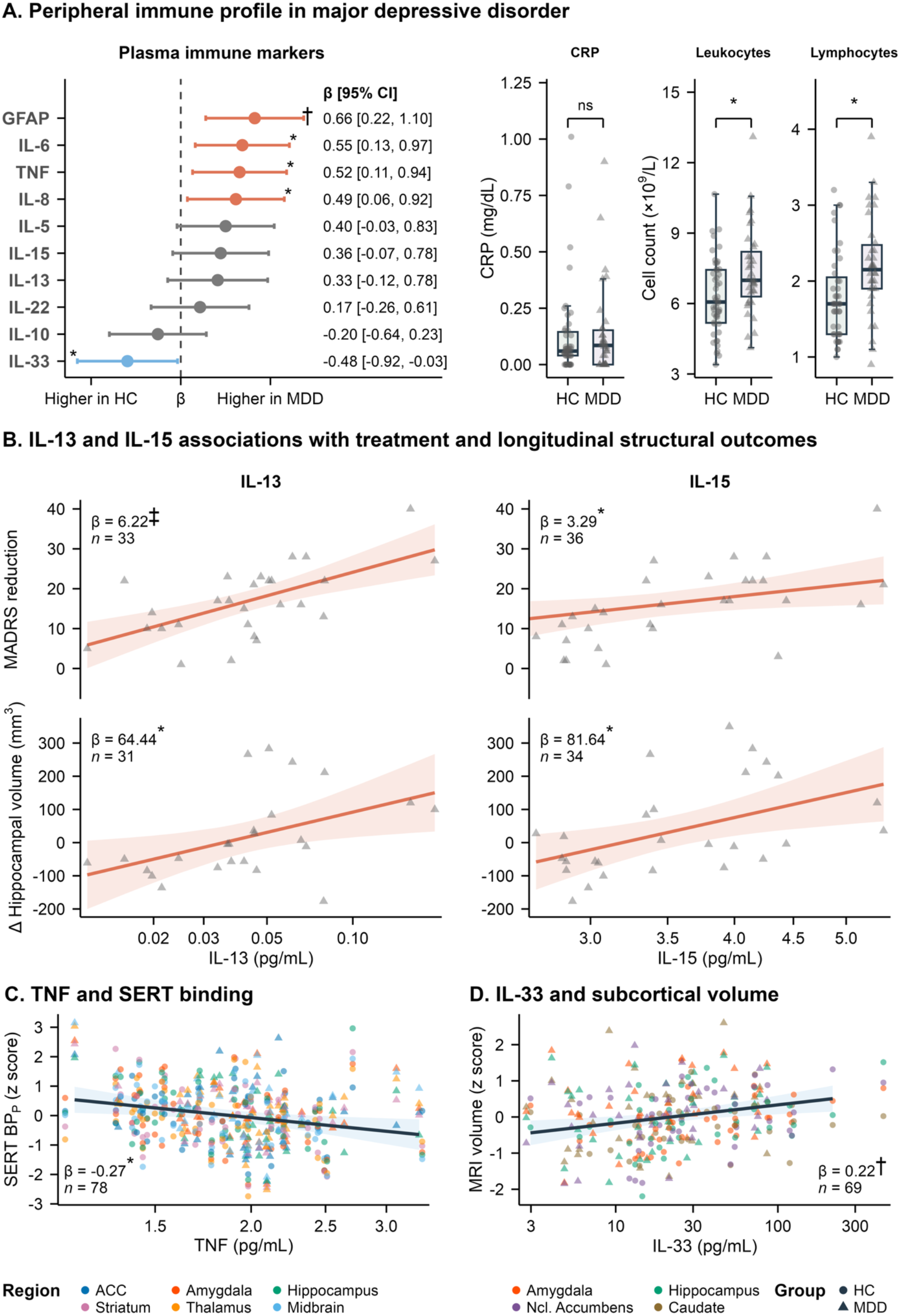
Detailed associations between plasma immune markers and clinical and neuroimaging outcomes. (A) Standardized differences in plasma immune-marker concentrations between patients with major depressive disorder (MDD) and healthy controls (HC), with positive coefficients indicating higher concentrations in MDD (left). Points show standardized regression coefficients and horizontal bars show 95% confidence intervals (CIs). Boxplots (right) show group differences in C-reactive protein (CRP) levels and leukocyte and lymphocyte counts. (B) Associations of baseline interleukin-13 (IL-13; left) and IL-15 (right) with improvement in Montgomery-Åsberg Depression Rating Scale (MADRS) score from baseline to week 6 (top) and change in hippocampal volume during treatment (bottom). Positive values indicate greater symptom improvement or increased hippocampal volume. (C) Association between baseline tumor necrosis factor (TNF) and regional serotonin transporter (SERT) binding at baseline. (D) Association between baseline IL-33 and subcortical brain volumes at baseline. Subcortical brain volumes are corrected for estimated total intracranial volume. In B-D, solid lines show model-estimated associations and shaded areas show 95% CIs. * nominal *p* < .05; † FDR-adjusted *p* < .05 within the corresponding domain; ‡ FDR-adjusted *p* < .05 across all analyses. ACC, anterior cingulate cortex; GFAP, glial fibrillary acidic protein; IL, interleukin.

Higher baseline IL-13 was associated with a greater improvement in depressive symptoms over six weeks of SSRI treatment. Each 1-SD increase in IL-13 corresponded to an additional 6.22-point reduction in MADRS score (β = 6.22, 95% CI 3.22 to 9.22, standardized β = 1.09, *p*_FDR,global_ = 0.01; Figure 2B). The association persisted after adjustment for covariates (β = 6.09, 95% CI 1.96 to 10.2). Exploratory analyses using HDRS_17_ (β = 4.04, 95% CI 1.71 to 6.37, *p* = 0.001) and BDI (β = 3.97, 95% CI 0.98 to 6.95, *p* = 0.01) showed concordant IL-13 associations. IL-15 was nominally associated with greater MADRS improvement (β = 3.29, 95% CI 0.27 to 6.31, *p* = 0.03).

No plasma marker was significantly associated with baseline symptom severity (all *p* ≥ 0.05).

Total leukocyte (β = 0.62, *p* < 0.01) and absolute lymphocyte counts (β = 0.64, *p* < 0.01) were higher in MDD patients, and these associations persisted after covariate adjustment (Figure 2A). No routine blood parameter was associated with antidepressant response, SERT binding, subcortical volume, cortical thickness, or hippocampal volume change with treatment (all p ≥ 0.1; Supplementary Table S10).

### Brain serotonergic transmission

No plasma marker was associated with *in vivo* SERT binding after FDR correction, and SERT binding did not differ by diagnosis (all *p* ≥ 0.45), as previously reported for this sample (Silberbauer et al., 2022). TNF showed a nominal inverse association with SERT binding across regions (β = −0.27, 95% CI −0.48 to −0.07, p = 0.01; Figure 2C). Adding sex as a covariate attenuated the estimate by 69% and adjustment for BMI by 26%, while other covariates had negligible influence.

### Brain structure

Baseline subcortical volume and cortical thickness did not differ significantly by diagnosis. Higher IL-33 was associated with greater baseline subcortical volume across the caudate, amygdala, hippocampus, and nucleus accumbens (β = 0.22, 95% CI 0.07 to 0.38, *p*_FDR,domain_ < 0.05; Figure 2D), without evidence that the association varied between regions (region × IL-33 interaction *p* > 0.22). This association was attenuated after controlling for covariates. (β = 0.16, 95% CI −0.00 to 0.32, *p* = 0.05).

Higher IL-15 was nominally associated with lower cortical thickness across the rostral and caudal anterior cingulate, insula, and medial orbitofrontal cortex (β = −0.23, 95% CI −0.39 to −0.06, *p* = 0.01, *p*_FDR,domain_ = 0.07). In the covariate-adjusted model, age alone reduced the IL-15 estimate by 61.4% (β = −0.05, *p* = 0.62) and was independently associated with lower cortical thickness (β = −0.05, 95% CI −0.07 to −0.03, *p* < 0.001).

Furthermore, higher IL-15 and IL-13 were nominally associated with greater longitudinal change in hippocampal volume during antidepressant treatment (IL-15: β = 81.64 mm^3^, 95% CI 19.34 to 143.93, *p* = 0.01; IL-13: β = 64.44 mm^3^, 95% CI 9.60 to 119.28, *p* = 0.02; Figure 2B). The IL-15 association was not markedly influenced by additional covariates or sensitivity analyses, but covariate adjustment reduced the IL-13 estimate. Moreover, greater six-week MADRS reduction correlated with more positive hippocampal volume change (ρ = 0.41, *p* = 0.04; Supplementary Figure S5).

## Discussion

In this multimodal study, we evaluated associations among high-sensitivity plasma immune markers, clinical characteristics, brain serotonergic transmission, and brain structure in individuals with MDD and healthy controls. Higher baseline IL-13 was associated with greater improvement in depressive symptoms during six weeks of SSRI treatment. This association survived correction across all primary tests, remained evident after covariate adjustment, and was directionally concordant across secondary symptom measures. Higher IL-15 showed a similar but nominal association with treatment response. Moreover, higher baseline IL-13 and IL-15 were both nominally associated with a more positive hippocampal volume change, although these associations did not survive FDR correction. GFAP concentrations were higher in MDD, and IL-33 was associated with greater subcortical volume at the domain-specific FDR level. No plasma marker was significantly associated with SERT binding or cortical thickness after multiple-testing correction. Baseline imaging measures did not differ by diagnosis, and diagnosis did not moderate immune-brain associations.

The higher GFAP concentrations in patients with MDD than in controls are consistent with reports of elevated serum and cerebrospinal GFAP, as well as increased GFAP in astrocyte-derived extracellular vesicles, in MDD and treatment-resistant depression (Michel et al., 2021; Steinacker et al., 2021; S. Xu et al., 2023). Nevertheless, treatment-naïve patients have also shown lower serum GFAP (Hviid et al., 2023), and a recent meta-analysis found no overall case-control difference (Kang et al., 2025). These inconsistencies may reflect differences in illness characteristics, treatment exposure, assay sensitivity, or the biological compartment examined. Circulating GFAP should also be interpreted as a nonspecific indicator of altered astroglial integrity or turnover rather than as direct evidence of astrogliosis or neurodegeneration. However, studies linking GFAP to astrocytic mitochondrial regulation and disrupted astrocyte-serotonin signaling to depression-related behavioral and synaptic abnormalities support the role of astroglial dysfunction in MDD (González-Arias et al., 2023; Xiong et al., 2025).

Nominal elevations of TNF, IL-6, and IL-8 were observed alongside increases in leukocyte counts, with both findings largely unchanged after covariate adjustment. Although these associations did not survive FDR correction, the direction of higher TNF and IL-6 aligns with meta-analytic findings in MDD, whereas evidence for IL-8 has been less consistent (Osimo et al., 2020). Recent work in first-episode, drug-naïve MDD identifies IL-6 rather than a uniform cytokine profile as the most reproducible signal (Zavaleta-Monestel et al., 2026). Lower IL-33 levels are also compatible with studies linking higher IL-33/ST2 concentrations to reduced depression risk. However, the context-dependent actions of this pathway preclude a straightforward protective interpretation (Li et al., 2025; Liu et al., 2023; Nahar et al., 2024). Collectively, these findings point to subtle and heterogeneous astroglial and immune alterations rather than a uniform inflammatory phenotype in MDD.

The strongest evidence concerned antidepressant response: higher IL-13 was associated with greater symptom improvement but not significantly related to baseline severity. IL-15 showed a similar nominal association, attenuated after covariate adjustment. Higher concentrations of both cytokines were also associated with greater increases in hippocampal volume during treatment. Importantly, greater improvement in MADRS was moderately correlated with a more positive change in hippocampal volume. The clinical and structural findings were therefore linked within individuals, rather than merely parallel, suggesting that IL-13 and IL-15 may index a broader treatment-responsiveness phenotype involving both symptomatic and hippocampal adaptation.

Previous findings support this paired signal: responders had higher pretreatment IL-13 and IL-15 concentrations than nonresponders, although neither cytokine alone showed strong discriminatory ability (Y. Xu et al., 2023). IL-13 was the most consistent moderator of depression outcome among 19 inflammatory markers, but its effect was treatment-specific. Higher IL-13 favored combination therapy with escitalopram and bupropion, whereas lower concentrations favored escitalopram monotherapy (Czysz et al., 2021). However, baseline IL-13 did not distinguish responders from nonresponders in other cohorts (Kofod et al., 2022; Schmidt et al., 2016), while meta-analytic evidence linked response to lower baseline IL-8 and greater treatment-related TNF reductions (Kofod et al., 2022; Köhler et al., 2018; Liu et al., 2020; Schmidt et al., 2016). Longitudinal findings were similarly heterogeneous. IL-13 decreased during treatment (Kofod et al., 2022; Schmidt et al., 2016) or shifted toward control levels (He et al., 2024), but its pooled change was nonsignificant (Köhler et al., 2018). IL-15 decreased transiently before returning to baseline in adolescents receiving fluoxetine (Pérez-Sánchez et al., 2018). Thus, the clinical relevance of IL-13 and IL-15 may depend on treatment context and whether baseline concentrations or treatment-related changes are examined.

Despite this clinical heterogeneity, both cytokines nevertheless have biological actions relevant to hippocampal plasticity. IL-13 and its receptors are present at neuronal synapses, where IL-13 can influence AMPA- and NMDA-receptor trafficking and excitatory transmission (Li et al., 2023). Signaling via the related IL-4Rα pathway also contributes to neuronal network homeostasis (Hanuscheck et al., 2022), and alterations in this pathway have been shown to change susceptibility to social stress (He et al., 2026). In models of stroke and traumatic brain injury, IL-13 promotes reparative microglial phenotypes and functional recovery (Chen et al., 2022; Kolosowska et al., 2019; Miao et al., 2020), although its effects are strongly context-dependent and can also aggravate neuronal injury and astrocyte loss (Hong et al., 2022). IL-15 regulates neural stem-cell proliferation and differentiation, while IL-15Rα supports hippocampal GABAergic and serotonergic function (He et al., 2010; Wu et al., 2011, 2010). Disruption of IL-15Rα signaling has also been associated with hippocampal microgliosis and astrogliosis, excessive synaptic pruning, and depression-like behavior (Tang et al., 2026). Together with evidence of treatment-related hippocampal plasticity and associations between decreasing inflammation and increasing hippocampal volume (Belge et al., 2020; Gryglewski et al., 2021; Phillips et al., 2015), the observed coupling between symptom improvement and changes in hippocampal volume during treatment motivates an immune-neuroplasticity hypothesis in which IL-13 and IL-15 may index biological states permissive of treatment adaptation.

Our PET findings provided little evidence for a relationship between peripheral immune variation and serotonergic transmission. TNF was nominally associated with lower SERT binding across regions, but covariate adjustment substantially reduced this estimate, suggesting possible demographic and metabolic confounding. Its direction also contrasts with the positive association between circulating TNF and brainstem SERT availability in a SPECT study (Krishnadas et al., 2016). Previous studies have linked SERT to IL-1-related cytokines or IL-6, although results have varied by population, region, and whether inflammatory activity was measured peripherally or centrally (Chou et al., 2016; Tsao et al., 2006; Yang et al., 2021). While TNF and IL-1β can experimentally enhance neuronal SERT activity (Zhu et al., 2006), transporter activity observed in cellular models does not directly correspond to *in vivo* radioligand binding. TNF may also affect serotonergic function independently of SERT by redirecting tryptophan metabolism toward the kynurenine pathway (Haroon et al., 2020; Okamoto et al., 2024). Thus, our data do not support a robust, generalized relationship between peripheral inflammation and SERT binding.

Higher IL-33 was associated with greater volume across the caudate, amygdala, hippocampus, and nucleus accumbens. Experimental studies show that neuronal IL-33/ST2 signaling supports synaptic remodeling in the hippocampus and integration of new neurons through microglial clearance of the extracellular matrix (Nguyen et al., 2020). In systemic-inflammatory and Alzheimer-like models, exogenous IL-33 also promotes reparative microglial responses and limits synaptic dysfunction, demyelination, and apoptosis (Fu et al., 2016; Nedeljkovic et al., 2026). These findings suggest a possible connection between neural tissue health and IL-33. However, peripheral inflammation is generally associated with reduced structural integrity in MDD, including smaller prefrontal grey matter, a thinner entorhinal cortex, and changes in hippocampal and amygdalar volumes (Green et al., 2021; Opel et al., 2019; Valizadeh et al., 2025). Higher IL-15 was nominally associated with lower thickness across the anterior cingulate, insula, and medial orbitofrontal cortex, broadly consistent with previous associations between peripheral inflammation and cortical alterations in MDD (Kakeda et al., 2018; Kang et al., 2024). However, this association appeared largely attributable to age-related confounding.

Strengths of this study involve combining high-sensitivity immune profiling with prospective clinical outcomes assessed at an unmedicated baseline, molecular imaging using [^11^C]DASB PET, and structural MRI within a single cohort. The MDD and healthy control groups were well matched. The young, physically healthy, normal-weight MDD cohort minimizes major confounders in peripheral immune analyses, including obesity, metabolic diseases, comorbidities, and concomitant medication. At the same time, this selectivity limits generalizability. Given the cohort’s mean age of 28.2 years, BMI of approximately 23, low CRP levels, and the absence of relevant somatic and psychiatric comorbidities, these findings may primarily reflect immune-brain relationships in a low-inflammatory MDD population, rather than in more heterogeneous clinical samples.

Additional limitations include small, modality-dependent sample sizes, and the absence of peripheral immune measurements at follow-up. Peripheral marker concentrations may not reflect signaling within the central nervous system, while PET binding and structural MRI remain indirect and biologically nonspecific measures. The observational design precludes causal inference, and the open-label treatment design cannot distinguish general prognostic associations from effects specific to antidepressant treatment.

Taken together, our findings support domain-specific immune associations instead of a unitary inflammatory signature of MDD. GFAP indicated astroglial alteration, IL-13 emerged as a candidate prognostic marker of treatment response, and IL-33 was associated with subcortical volume. Overall, peripheral immune variation appeared more closely related to dynamic treatment phenotypes than to static serotonergic or structural brain measures, as further suggested by the exploratory convergence of IL-13 and IL-15 across changes in symptoms and hippocampal volume. These findings provide a focused framework for prospective studies testing whether peripheral immune profiles can predict and potentially shape clinical and structural adaptation during antidepressant treatment.

## Data Availability

All data produced in the present study are available upon reasonable request to the authors

## ClinicalTrials.gov

Patient Stratification and Treatment Response Prediction in Neuropharmacotherapy Using Hybrid Positron Emission Tomography/Magnetic Resonance Imaging (PET/MR); https://clinicaltrials.gov/study/NCT02711215; NCT02711215.

## Abbreviations

ACC: anterior cingulate cortex
BDI: Beck Depression Inventory
BMI: body mass index
BP_P_: binding potential
CRP: C-reactive protein
eTIV: estimated total intracranial volume
GFAP: glial fibrillary acidic protein
GPAQ: Global Physical Activity Questionnaire
HC: healthy controls
HDRS_17_: 17-item Hamilton Depression Rating Scale
IL: interleukin
MADRS: Montgomery-Åsberg Depression Rating Scale
MDD: major depressive disorder
MRI: magnetic resonance imaging
PET: positron emission tomography
SERT: serotonin transporter
Simoa: single-molecule array
SSRI: selective serotonin reuptake inhibitor
TNF: tumor necrosis factor

## Acknowledgements

We thank all the participants who took part in this study. Our appreciation extends to Marius Hienert, Thomas Vanicek, Alexander Kautzky, Johannes Jungwirth, Paul Michenthaler, Alim Emre Başaran, Radheshyam Stepponat, and the medical students of the Neuroimaging Labs for their medical support; to Edda Winkler-Pjrek, Siegfried Kasper, and Markus Hartenbach for medical supervision; and to Georg Kranz for assisting with participant randomization. We also thank Karoline Einenkel, Elisa Sittenberger, and Vera Ritter for administrative support, and Benjamin Spurny, Murray B. Reed and Gregory M. James for technical support. We gratefully acknowledge the contributions of Verena Pichler, Neydher Berroterán-Infante, Theresa Balber, Markus Mitterhauser, Wolfgang Wadsak, and Eva-Maria Klebermass to tracer synthesis, metabolite analysis, and supervision.

## Author contributions

**Benjamin Eggerstorfer**: Methodology, Software, Validation, Formal analysis, Investigation, Data curation, Writing – original draft, Visualization. **Patrick Altmann**: Methodology, Formal analysis. **Matej Murgaš**: Software, Formal analysis, Investigation. **Markus Ponleitner**: Investigation. **Godber M. Godbersen**: Investigation. **Manfred Klöbl**: Investigation. **Jakob Unterholzner**: Investigation. **Leo R. Silberbauer**: Investigation, Data curation. **Chrysoula Vraka**: Investigation. **Cecile Philippe**: Investigation. **Lukas Nics**: Resources, Supervision. **Andreas Hahn**: Resources, Supervision. **Marcus Hacker**: Resources, Supervision. **Gregor Gryglewski**: Conceptualization, Methodology, Software, Validation, Formal analysis, Investigation, Data curation, Writing – original draft, Supervision, Project administration. **Rupert Lanzenberger**: Conceptualization, Resources, Supervision, Project administration, Funding acquisition. **All authors**: Writing – review & editing.

## Declaration of generative AI and AI-assisted technologies in the manuscript preparation process

During the preparation of this work the author(s) used ChatGPT (OpenAI) in order to assist with generating R code for data visualization and with grammar and language editing. After using this tool/service, the author(s) reviewed and edited the content as needed and take(s) full responsibility for the content of the published article.

