## Supplement for "Peripheral immune biomarkers, brain imaging phenotypes and treatment response in major depressive disorder"

### Supplementary Methods

#### Single-molecule array quality control

Interleukins (IL-5, IL-6, IL-8, IL-10, IL-13, IL-15, IL-17A, IL-22, IL-33), TNF, and GFAP were included in analyses. Considering the log-normal distribution and the multiplicative error structure, replicate agreement was assessed on the log scale. Pairs with a geometric coefficient of variation exceeding 20% were omitted (Supplementary Figure S2), and geometric means were used for subsequent analysis.

Immune markers underwent quality control according to their detection frequency to manage left-censoring. Those detected in 80% or more of samples were analyzed as continuous variables after log transformation. Markers detected in 20–80% of samples were evaluated as binary, while markers with less than 20% detection were omitted from quantitative analysis.

#### Single-molecule array analyses consumables

The following Quanterix Simoa assay kits were utilized: Cytokine 3-Plex A assay (catalog no. 101160), GFAP Simoa Discovery Kit (catalog no. 102336), IL-17A Simoa 2.0 Assay Kit (catalog no. 101599), IL-5 Simoa Assay Kit (catalog no. 102860), IL-8 Simoa Assay Kit (catalog no. 100198), IL-13 Simoa Assay Kit (catalog no. 102732), IL-15 Simoa Assay Kit (catalog no. 100794), IL-22 (Total) Simoa Discovery Assay Kit (catalog no. 103071), and IL-33 Simoa Discovery Kit (catalog no. 103093). SR-X consumables and reagents included the SR-X Consumables Kit (catalog no. 03077), Wash Buffer A (catalog no. 03078), Wash Buffer B (catalog no. 103079), and Sealing Oil for SR-X (catalog no. 102767).

#### PET-imaging procedures and quantification of serotonin transporter (SERT) binding potentials

PET/MR acquisition and quantification of serotonin transporter (SERT) binding were carried out using a well-established [$^{11}$C]DASB bolus plus constant infusion method (Gryglewski et al., 2017). Participants completed two PET/MR sessions in a randomized, placebo-controlled crossover design, with at least 3 days between each session. In one session, they received 8 mg of citalopram intravenously; in the other, saline was administered. This analysis included only data from the placebo session's imaging and plasma samples.

On each scanning day, arterial and venous catheters were placed. The SERT-selective radioligand [$^{11}$C]DASB ([$^{11}$C]-N,N-dimethyl-2-(2-amino-4-cyanophenylthio)benzylamine) was administered as a bolus, then followed by constant infusion. During the placebo session, saline was infused over 8 minutes starting 70 minutes after radioligand administration began. Arterial blood was sampled using 9-mL lithium-heparin S-Monovettes (92 × 16 mm; catalog no. 02.1065, Sarstedt) at predefined time points for measurement of metabolite-corrected plasma radioactivity.

PET data were acquired in list mode on an integrated Biograph mMR PET/MR system (Siemens Healthineers, Erlangen, Germany) and reconstructed using an ordinary Poisson ordered-subsets expectation-maximization algorithm with three iterations and 21 subsets. Attenuation correction utilized a low-dose CT scan obtained on a different occasion with a Siemens Biograph TruePoint PET/CT system.

PET images were coregistered to a T1-weighted structural image acquired using an MPRAGE sequence (TE/TR = 4.21/2000 ms; in-plane resolution, 1 × 1 mm; slice thickness, 1 mm; interslice gap, 0.1 mm) and spatially normalized to Montreal Neurological Institute space using SPM12 (Wellcome Centre for Human Neuroimaging, London, UK). Regions of interest were defined using the Automated Anatomical Labeling atlas.

At equilibrium, regional tissue radioactivity was averaged over the predefined analysis interval and divided by the mean metabolite-corrected plasma radioactivity to estimate the total distribution volume, $V_T$. The plasma-reference binding potential, $BP_P$, was calculated as

$$BP_P = V_T - V_{ND}$$

where $V_{ND}$ was approximated using the distribution volume of cerebellar grey matter, chosen as the reference region due to its low specific SERT binding.

**Structural magnetic resonance imaging and image preprocessing**

Structural MRI data were acquired using an integrated 3-T Biograph mMR system (Siemens Healthineers, Erlangen, Germany). Baseline MRI data were collected during the PET/MR sessions, and the mean of the regional estimate across both sessions was computed for further analyses. Follow-up MRI data were acquired on the same scanner in an MRI-only session without simultaneous PET acquisition. A T1-weighted MPRAGE sequence was performed with an echo time/repetition time of 4.21/2000 ms, an in-plane resolution of 1 × 1 mm, a slice thickness of 1 mm, and an interslice gap of 0.1 mm.

Structural images were processed using FreeSurfer version 6.0. Cortical reconstruction and automated subcortical segmentation were performed using the standard FreeSurfer processing pipeline. All segmentations were visually inspected for major processing errors and corrected where necessary. Regional volumes were extracted from the resulting anatomical parcellations. Estimated total intracranial volume was calculated using FreeSurfer to account for variations in head size among individuals.

Baseline and follow-up images were processed using the FreeSurfer longitudinal pipeline.

**Model diagnostics**

Model diagnostics included convergence and singularity assessments, residual-versus-fitted plots, residual Q-Q plots, and participant-level influence screening. Findings reaching nominal or FDR-adjusted significance were examined by leave-one-participant-out refitting to assess coefficient stability and sign reversals. For regional analyses, leave-one-region-out models assessed whether common-slope findings depended on any individual regions.

### Supplementary Tables

**Supplementary Table S1: Single-molecule array replicates quality control summary table.** For each marker, the table reports the total number of replicates analyzed, numbers and percentages below the lower limit of detection (LOD), imputed, excluded because of high geometric coefficient of variation (gCV), and retained after quality control, together with the median gCV and assay-specific LOD and lower limit of quantification (LLOQ).

| Marker | n (total) | n (below LOD) | n (imputed) | n (high gCV) | n (kept replicate) | Median gCV | n (below LOD, %) | n (imputed, %) | N (high gCV, %) | N (kept replicate, %) | LOD (pg/mL) | LLOQ (pg/mL) |
|---|---|---|---|---|---|---|---|---|---|---|---|---|
| GFAP | 178 | 2 | 0 | 16 | 160 | 5.64 | 1.1 | 0 | 9 | 89.9 | 0.26 | 1.37 |
| IL-10 | 178 | 0 | 0 | 0 | 177 | 3.13 | 0 | 0 | 0 | 99.4 | 0.0022 | 0.0073 |
| IL-13 | 178 | 0 | 0 | 14 | 164 | 6.62 | 0 | 0 | 7.9 | 92.1 | 0.001 | 0.007 |
| IL-15 | 178 | 0 | 0 | 0 | 177 | 3.30 | 0 | 0 | 0 | 99.4 | 0.002 | 0.0069 |
| IL-17A | 178 | 14 | 0 | 24 | 128 | 7.84 | 7.9 | 0 | 13.5 | 71.9 | 0.011 | 0.011 |
| IL-22 | 178 | 0 | 0 | 2 | 173 | 3.79 | 0 | 0 | 1.1 | 97.2 | 0.004 | 0.021 |
| IL-33 | 178 | 0 | 0 | 26 | 152 | 8.17 | 0 | 0 | 14.6 | 85.4 | 0.32 | 0.686 |
| IL-5 | 178 | 0 | 0 | 8 | 167 | 5.71 | 0 | 0 | 4.5 | 93.8 | 0.0034 | 0.0165 |
| IL-6 | 178 | 0 | 0 | 2 | 175 | 5.78 | 0 | 0 | 1.1 | 98.3 | 0.006 | 0.011 |
| IL-8 | 178 | 0 | 0 | 8 | 168 | 4.87 | 0 | 0 | 4.5 | 94.4 | 0.038 | 0.146 |
| TNF | 178 | 0 | 0 | 0 | 177 | 4.14 | 0 | 0 | 0 | 99.4 | 0.011 | 0.051 |

**Supplementary Table S2: Full results of associations between immune markers and clinical and neuroimaging outcomes.** Associations between each immune marker and diagnosis, antidepressant response, baseline serotonin transporter (SERT) binding, baseline subcortical volume, baseline cortical thickness, and treatment-related hippocampal volume change. Longitudinal models tested the timepoint × immune marker interaction. p, nominal p; $p_{FDR,domain}$, FDR-adjusted within the corresponding domain; $p_{FDR,global}$, FDR-adjusted across all primary analyses. LMM, linear mixed-effects model; MADRS, Montgomery-Åsberg Depression Rating Scale; SERT, serotonin transporter; $BP_P$, binding potential relative to plasma; eTIV, estimated total intracranial volume; GFAP, glial fibrillary acidic protein; IL, interleukin; TNF, tumor necrosis factor.

| **Domain** | **Marker** | **n** | **Model** | **β** | **95% CI** | **statistic** | **p** | **$p_{FDR,domain}$** | **$p_{FDR,global}$** | **Signed z** |
|---|---|---|---|---|---|---|---|---|---|---|
| Diagnosis | GFAP | 75 | immune marker ~ group | 0.66 | [0.215, 1.105] | 2.96 | 0.004 | 0.042 | 0.100 | 2.87 |
| Diagnosis | IL-6 | 83 | immune marker ~ group | 0.55 | [0.123, 0.976] | 2.56 | 0.012 | 0.055 | 0.105 | 2.51 |
| Diagnosis | TNF | 84 | immune marker ~ group | 0.53 | [0.099, 0.951] | 2.45 | 0.016 | 0.055 | 0.123 | 2.40 |
| Diagnosis | IL-8 | 80 | immune marker ~ group | 0.49 | [0.052, 0.931] | 2.23 | 0.029 | 0.072 | 0.173 | 2.19 |
| Diagnosis | IL-33 | 74 | immune marker ~ group | -0.48 | [-0.93, -0.021] | -2.09 | 0.041 | 0.081 | 0.203 | -2.05 |
| Diagnosis | IL-5 | 81 | immune marker ~ group | 0.40 | [-0.04, 0.841] | 1.81 | 0.074 | 0.123 | 0.273 | 1.79 |
| Diagnosis | IL-15 | 84 | immune marker ~ group | 0.36 | [-0.078, 0.791] | 1.63 | 0.107 | 0.152 | 0.304 | 1.61 |
| Diagnosis | IL-13 | 78 | immune marker ~ group | 0.33 | [-0.123, 0.783] | 1.45 | 0.151 | 0.189 | 0.336 | 1.44 |
| Diagnosis | IL-10 | 84 | immune marker ~ group | -0.20 | [-0.643, 0.235] | -0.93 | 0.357 | 0.397 | 0.596 | -0.92 |
| Diagnosis | IL-22 | 83 | immune marker ~ group | 0.17 | [-0.271, 0.616] | 0.77 | 0.442 | 0.442 | 0.631 | 0.77 |
| Antidepressant response | IL-13 | 33 | LMM: MADRS ~ timepoint * immune marker + (1 \| subject) | 6.22 | [3.219, 9.218] | 4.22 | 0.000 | 0.002 | 0.011 | 3.74 |
| Antidepressant response | IL-15 | 36 | LMM: MADRS ~ timepoint * immune marker + (1 \| subject) | 3.29 | [0.268, 6.306] | 2.22 | 0.034 | 0.169 | 0.184 | 2.12 |
| Antidepressant response | IL-5 | 35 | LMM: MADRS ~ timepoint * immune marker + (1 \| subject) | 2.93 | [-0.44, 6.305] | 1.76 | 0.086 | 0.267 | 0.273 | 1.72 |
| Antidepressant response | TNF | 36 | LMM: MADRS ~ timepoint * immune marker + (1 \| subject) | -2.89 | [-6.566, 0.792] | -1.59 | 0.120 | 0.267 | 0.310 | -1.55 |

| | | | | | | | | | | |
|---|---|---|---|---|---|---|---|---|---|---|
| Antidepressant response | IL-33 | 34 | LMM: MADRS ~ timepoint * immune marker + (1 \| subject) | -2.70 | [-6.274, 0.876] | -1.54 | 0.134 | 0.267 | 0.310 | -1.50 |
| Antidepressant response | GFAP | 31 | LMM: MADRS ~ timepoint * immune marker + (1 \| subject) | -3.02 | [-8.152, 2.12] | -1.20 | 0.239 | 0.398 | 0.469 | -1.18 |
| Antidepressant response | IL-8 | 34 | LMM: MADRS ~ timepoint * immune marker + (1 \| subject) | 2.13 | [-2.196, 6.452] | 1.01 | 0.323 | 0.461 | 0.570 | 0.99 |
| Antidepressant response | IL-10 | 36 | LMM: MADRS ~ timepoint * immune marker + (1 \| subject) | -1.84 | [-6.209, 2.532] | -0.85 | 0.400 | 0.500 | 0.623 | -0.84 |
| Antidepressant response | IL-6 | 36 | LMM: MADRS ~ timepoint * immune marker + (1 \| subject) | 0.73 | [-2.366, 3.825] | 0.48 | 0.635 | 0.705 | 0.747 | 0.48 |
| Antidepressant response | IL-22 | 35 | LMM: MADRS ~ timepoint * immune marker + (1 \| subject) | 0.09 | [-4.856, 5.037] | 0.04 | 0.971 | 0.971 | 0.995 | 0.04 |
| Baseline SERT binding | TNF | 78 | LMM: SERT $BP_P$ ~ immune marker + region + group + (1 \| subject) | -0.27 | [-0.475, -0.065] | -2.63 | 0.011 | 0.105 | 0.105 | -2.56 |
| Baseline SERT binding | IL-5 | 75 | LMM: SERT $BP_P$ ~ immune marker + region + group + (1 \| subject) | 0.20 | [-0.015, 0.405] | 1.85 | 0.069 | 0.280 | 0.273 | 1.82 |
| Baseline SERT binding | IL-8 | 74 | LMM: SERT $BP_P$ ~ immune marker + region + group + (1 \| subject) | -0.18 | [-0.389, 0.025] | -1.75 | 0.084 | 0.280 | 0.273 | -1.73 |
| Baseline SERT binding | IL-13 | 72 | LMM: SERT $BP_P$ ~ immune marker + region + group + (1 \| subject) | 0.13 | [-0.091, 0.359] | 1.19 | 0.237 | 0.594 | 0.469 | 1.18 |
| Baseline SERT binding | IL-6 | 77 | LMM: SERT $BP_P$ ~ immune marker + region + group + (1 \| subject) | 0.09 | [-0.113, 0.291] | 0.88 | 0.383 | 0.738 | 0.621 | 0.87 |
| Baseline SERT binding | IL-22 | 77 | LMM: SERT $BP_P$ ~ immune marker + region + group + (1 \| subject) | -0.06 | [-0.261, 0.141] | -0.60 | 0.552 | 0.738 | 0.712 | -0.60 |
| Baseline SERT binding | IL-10 | 78 | LMM: SERT $BP_P$ ~ immune marker + region + group + (1 \| subject) | -0.06 | [-0.268, 0.146] | -0.59 | 0.556 | 0.738 | 0.712 | -0.59 |
| Baseline SERT binding | IL-15 | 78 | LMM: SERT $BP_P$ ~ immune marker + region + group + (1 \| subject) | 0.05 | [-0.146, 0.254] | 0.54 | 0.591 | 0.738 | 0.723 | 0.54 |
| Baseline SERT binding | IL-33 | 68 | LMM: SERT $BP_P$ ~ immune marker + region + group + (1 \| subject) | -0.02 | [-0.244, 0.203] | -0.18 | 0.856 | 0.884 | 0.947 | -0.18 |
| Baseline SERT binding | GFAP | 70 | LMM: SERT $BP_P$ ~ immune marker + region + group + (1 \| subject) | 0.02 | [-0.217, 0.251] | 0.15 | 0.884 | 0.884 | 0.947 | 0.15 |
| Baseline subcortical volume | IL-33 | 69 | LMM: subcortical volume ~ immune marker + region + eTIV + group + (1 \| subject) | 0.22 | [0.07, 0.378] | 2.91 | 0.0048 | 0.0498 | 0.100 | 2.81 |

| | | | | | | | | | | |
|---|---|---|---|---|---|---|---|---|---|---|
| Baseline subcortical volume | IL-6 | 77 | LMM: subcortical volume ~ immune marker + region + eTIV + group + (1 \| subject) | -0.14 | [-0.299, 0.011] | -1.85 | 0.069 | 0.254 | 0.273 | -1.82 |
| Baseline subcortical volume | IL-5 | 75 | LMM: subcortical volume ~ immune marker + region + eTIV + group + (1 \| subject) | -0.14 | [-0.307, 0.02] | -1.75 | 0.085 | 0.254 | 0.273 | -1.72 |
| Baseline subcortical volume | IL-15 | 78 | LMM: subcortical volume ~ immune marker + region + eTIV + group + (1 \| subject) | -0.12 | [-0.271, 0.025] | -1.66 | 0.102 | 0.254 | 0.304 | -1.64 |
| Baseline subcortical volume | IL-13 | 72 | LMM: subcortical volume ~ immune marker + region + eTIV + group + (1 \| subject) | -0.13 | [-0.296, 0.039] | -1.54 | 0.129 | 0.259 | 0.310 | -1.52 |
| Baseline subcortical volume | IL-8 | 74 | LMM: subcortical volume ~ immune marker + region + eTIV + group + (1 \| subject) | -0.09 | [-0.253, 0.065] | -1.18 | 0.242 | 0.404 | 0.469 | -1.17 |
| Baseline subcortical volume | IL-10 | 78 | LMM: subcortical volume ~ immune marker + region + eTIV + group + (1 \| subject) | -0.06 | [-0.219, 0.098] | -0.76 | 0.452 | 0.646 | 0.631 | -0.75 |
| Baseline subcortical volume | GFAP | 70 | LMM: subcortical volume ~ immune marker + region + eTIV + group + (1 \| subject) | 0.05 | [-0.119, 0.227] | 0.62 | 0.537 | 0.671 | 0.712 | 0.62 |
| Baseline subcortical volume | TNF | 78 | LMM: subcortical volume ~ immune marker + region + eTIV + group + (1 \| subject) | 0.02 | [-0.149, 0.18] | 0.19 | 0.853 | 0.948 | 0.947 | 0.19 |
| Baseline subcortical volume | IL-22 | 77 | LMM: subcortical volume ~ immune marker + region + eTIV + group + (1 \| subject) | 0.00 | [-0.157, 0.158] | 0.01 | 0.995 | 0.995 | 0.995 | 0.01 |
| Baseline cortical thickness | IL-15 | 78 | LMM: cortical thickness ~ immune marker + region + group + (1 \| subject) | -0.23 | [-0.392, -0.064] | -2.77 | 0.007 | 0.071 | 0.105 | -2.69 |
| Baseline cortical thickness | GFAP | 70 | LMM: cortical thickness ~ immune marker + region + group + (1 \| subject) | -0.17 | [-0.361, 0.019] | -1.79 | 0.077 | 0.336 | 0.273 | -1.77 |
| Baseline cortical thickness | IL-5 | 75 | LMM: cortical thickness ~ immune marker + region + group + (1 \| subject) | -0.15 | [-0.33, 0.039] | -1.57 | 0.121 | 0.336 | 0.310 | -1.55 |
| Baseline cortical thickness | IL-6 | 77 | LMM: cortical thickness ~ immune marker + region + group + (1 \| subject) | -0.13 | [-0.306, 0.042] | -1.51 | 0.134 | 0.336 | 0.310 | -1.50 |
| Baseline cortical thickness | IL-8 | 74 | LMM: cortical thickness ~ immune marker + region + group + (1 \| subject) | -0.10 | [-0.279, 0.083] | -1.08 | 0.285 | 0.571 | 0.535 | -1.07 |

| | | | | | | | | | | |
|---|---|---|---|---|---|---|---|---|---|---|
| Baseline cortical thickness | IL-13 | 72 | LMM: cortical thickness ~ immune marker + region + group + (1 \| subject) | -0.08 | [-0.272, 0.112] | -0.83 | 0.409 | 0.681 | 0.623 | -0.83 |
| Baseline cortical thickness | TNF | 78 | LMM: cortical thickness ~ immune marker + region + group + (1 \| subject) | -0.06 | [-0.243, 0.132] | -0.59 | 0.557 | 0.713 | 0.712 | -0.59 |
| Baseline cortical thickness | IL-33 | 69 | LMM: cortical thickness ~ immune marker + region + group + (1 \| subject) | 0.05 | [-0.134, 0.242] | 0.57 | 0.571 | 0.713 | 0.713 | 0.57 |
| Baseline cortical thickness | IL-10 | 78 | LMM: cortical thickness ~ immune marker + region + group + (1 \| subject) | -0.03 | [-0.215, 0.146] | -0.38 | 0.707 | 0.785 | 0.816 | -0.38 |
| Baseline cortical thickness | IL-22 | 77 | LMM: cortical thickness ~ immune marker + region + group + (1 \| subject) | -0.02 | [-0.195, 0.166] | -0.16 | 0.873 | 0.873 | 0.947 | -0.16 |
| Hippocampal volume change with treatment | IL-15 | 34 | LMM: hippocampal volume ~ timepoint * immune marker + eTIV + (1 \| subject) | 81.64 | [19.344, 143.934] | 2.70 | 0.012 | 0.117 | 0.105 | 2.50 |
| Hippocampal volume change with treatment | IL-13 | 31 | LMM: hippocampal volume ~ timepoint * immune marker + eTIV + (1 \| subject) | 64.44 | [9.603, 119.281] | 2.44 | 0.023 | 0.117 | 0.156 | 2.27 |
| Hippocampal volume change with treatment | IL-8 | 32 | LMM: hippocampal volume ~ timepoint * immune marker + eTIV + (1 \| subject) | 46.49 | [-23.442, 116.421] | 1.38 | 0.182 | 0.608 | 0.391 | 1.33 |
| Hippocampal volume change with treatment | IL-5 | 33 | LMM: hippocampal volume ~ timepoint * immune marker + eTIV + (1 \| subject) | 32.33 | [-32.27, 96.936] | 1.03 | 0.312 | 0.609 | 0.567 | 1.01 |
| Hippocampal volume change with treatment | IL-22 | 33 | LMM: hippocampal volume ~ timepoint * immune marker + eTIV + (1 \| subject) | -40.05 | [-123.676, 43.572] | -0.99 | 0.333 | 0.609 | 0.572 | -0.97 |
| Hippocampal volume change with treatment | IL-6 | 34 | LMM: hippocampal volume ~ timepoint * immune marker + eTIV + (1 \| subject) | 20.23 | [-30.261, 70.723] | 0.83 | 0.417 | 0.609 | 0.623 | 0.81 |
| Hippocampal volume change with treatment | GFAP | 30 | LMM: hippocampal volume ~ timepoint * immune marker + eTIV + (1 \| subject) | -31.38 | [-111.589, 48.833] | -0.81 | 0.426 | 0.609 | 0.623 | -0.80 |
| Hippocampal volume change with treatment | TNF | 34 | LMM: hippocampal volume ~ timepoint * immune marker + eTIV + (1 \| subject) | -16.58 | [-83.748, 50.59] | -0.51 | 0.616 | 0.770 | 0.739 | -0.50 |
| Hippocampal volume change with treatment | IL-33 | 32 | LMM: hippocampal volume ~ timepoint * immune marker + eTIV + (1 \| subject) | 2.87 | [-63.008, 68.745] | 0.09 | 0.929 | 0.979 | 0.978 | 0.09 |
| Hippocampal volume change with treatment | IL-10 | 34 | LMM: hippocampal volume ~ timepoint * immune marker + eTIV + (1 \| subject) | -1.09 | [-86.194, 84.024] | -0.03 | 0.979 | 0.979 | 0.995 | -0.03 |

**Supplementary Table S3: Exploratory covariate-adjusted associations between immune markers and clinical and neuroimaging outcomes.** Models were adjusted for age, sex, body mass index (BMI), and total score of Global Physical Activity Questionnaire (GPAQ). Longitudinal models tested the timepoint × immune marker interaction. p, nominal p; $p_{FDR,domain}$, FDR-adjusted within the corresponding domain. LMM, linear mixed effects model; MADRS, Montgomery-Åsberg Depression Rating Scale; SERT, serotonin transporter; $BP_P$, binding potential relative to plasma; GFAP, glial fibrillary acidic protein; IL, interleukin; TNF, tumor necrosis factor.

| Domain | Marker | n | Model | β | 95% CI | Statistic | p | $p_{FDR,domain}$ |
|---|---|---|---|---|---|---|---|---|
| Diagnosis | IL-8 | 67 | immune marker ~ group + age + sex + BMI + GPAQ total | 0.66 | [0.236, 1.081] | 3.12 | 0.003 | 0.021 |
| Diagnosis | GFAP | 61 | immune marker ~ group + age + sex + BMI + GPAQ total | 0.78 | [0.257, 1.3] | 2.991 | 0.004 | 0.021 |
| Diagnosis | IL-5 | 67 | immune marker ~ group + age + sex + BMI + GPAQ total | 0.58 | [0.079, 1.084] | 2.314 | 0.024 | 0.066 |
| Diagnosis | IL-6 | 68 | immune marker ~ group + age + sex + BMI + GPAQ total | 0.57 | [0.07, 1.078] | 2.278 | 0.026 | 0.066 |
| Diagnosis | TNF | 69 | immune marker ~ group + age + sex + BMI + GPAQ total | 0.48 | [0.022, 0.94] | 2.092 | 0.041 | 0.081 |
| Diagnosis | IL-15 | 69 | immune marker ~ group + age + sex + BMI + GPAQ total | 0.44 | [-0.021, 0.897] | 1.908 | 0.061 | 0.101 |
| Diagnosis | IL-33 | 60 | immune marker ~ group + age + sex + BMI + GPAQ total | -0.44 | [-0.994, 0.111] | -1.601 | 0.115 | 0.149 |
| Diagnosis | IL-13 | 63 | immune marker ~ group + age + sex + BMI + GPAQ total | 0.44 | [-0.117, 1] | 1.583 | 0.119 | 0.149 |
| Diagnosis | IL-22 | 68 | immune marker ~ group + age + sex + BMI + GPAQ total | 0.14 | [-0.418, 0.7] | 0.504 | 0.616 | 0.684 |
| Diagnosis | IL-10 | 69 | immune marker ~ group + age + sex + BMI + GPAQ total | -0.02 | [-0.343, 0.309] | -0.105 | 0.917 | 0.917 |
| Antidepressant response | IL-13 | 22 | LMM: MADRS ~ timepoint * immune marker + age + sex + BMI + GPAQ total + (1 \| subject) | 6.09 | [1.963, 10.21] | 3.071 | 0.006 | 0.058 |
| Antidepressant response | IL-10 | 25 | LMM: MADRS ~ timepoint * immune marker + age + sex + BMI + GPAQ total + (1 \| subject) | -5.61 | [-10.753, -0.475] | -2.266 | 0.034 | 0.168 |
| Antidepressant response | IL-33 | 23 | LMM: MADRS ~ timepoint * immune marker + age + sex + BMI + GPAQ total + (1 \| subject) | -3.42 | [-8.084, 1.254] | -1.528 | 0.143 | 0.301 |
| Antidepressant response | IL-15 | 25 | LMM: MADRS ~ timepoint * immune marker + age + sex + BMI + GPAQ total + (1 \| subject) | 2.75 | [-1.051, 6.55] | 1.505 | 0.147 | 0.301 |
| Antidepressant response | TNF | 25 | LMM: MADRS ~ timepoint * immune marker + age + sex + BMI + GPAQ total + (1 \| subject) | -3.54 | [-8.466, 1.391] | -1.489 | 0.151 | 0.301 |

| | | | | | | | | |
|---|---|---|---|---|---|---|---|---|
| Antidepressant response | GFAP | 21 | LMM: MADRS ~ timepoint * immune marker + age + sex + BMI + GPAQ total + (1 \| subject) | -5.20 | [-13.268, 2.869] | -1.349 | 0.193 | 0.307 |
| Antidepressant response | IL-5 | 25 | LMM: MADRS ~ timepoint * immune marker + age + sex + BMI + GPAQ total + (1 \| subject) | 2.70 | [-1.666, 7.057] | 1.271 | 0.215 | 0.307 |
| Antidepressant response | IL-22 | 24 | LMM: MADRS ~ timepoint * immune marker + age + sex + BMI + GPAQ total + (1 \| subject) | -0.75 | [-6.905, 5.403] | -0.255 | 0.801 | 0.993 |
| Antidepressant response | IL-6 | 25 | LMM: MADRS ~ timepoint * immune marker + age + sex + BMI + GPAQ total + (1 \| subject) | -0.02 | [-3.516, 3.468] | -0.014 | 0.989 | 0.993 |
| Antidepressant response | IL-8 | 23 | LMM: MADRS ~ timepoint * immune marker + age + sex + BMI + GPAQ total + (1 \| subject) | -0.03 | [-7.312, 7.25] | -0.009 | 0.993 | 0.993 |
| Baseline SERT binding | IL-5 | 64 | LMM: SERT BPP ~ immune marker + region + group + age + sex + BMI + GPAQ total + (1 \| subject) | 0.23 | [0.046, 0.422] | 2.494 | 0.016 | 0.156 |
| Baseline SERT binding | GFAP | 58 | LMM: SERT BPP ~ immune marker + region + group + age + sex + BMI + GPAQ total + (1 \| subject) | -0.13 | [-0.35, 0.089] | -1.194 | 0.238 | 0.814 |
| Baseline SERT binding | IL-13 | 60 | LMM: SERT BPP ~ immune marker + region + group + age + sex + BMI + GPAQ total + (1 \| subject) | 0.12 | [-0.081, 0.312] | 1.178 | 0.244 | 0.814 |
| Baseline SERT binding | IL-8 | 64 | LMM: SERT BPP ~ immune marker + region + group + age + sex + BMI + GPAQ total + (1 \| subject) | -0.11 | [-0.345, 0.117] | -0.989 | 0.327 | 0.817 |
| Baseline SERT binding | TNF | 66 | LMM: SERT BPP ~ immune marker + region + group + age + sex + BMI + GPAQ total + (1 \| subject) | -0.07 | [-0.277, 0.138] | -0.666 | 0.508 | 0.897 |
| Baseline SERT binding | IL-6 | 65 | LMM: SERT BPP ~ immune marker + region + group + age + sex + BMI + GPAQ total + (1 \| subject) | -0.05 | [-0.236, 0.14] | -0.512 | 0.611 | 0.897 |
| Baseline SERT binding | IL-10 | 66 | LMM: SERT BPP ~ immune marker + region + group + age + sex + BMI + GPAQ total + (1 \| subject) | 0.05 | [-0.236, 0.342] | 0.364 | 0.717 | 0.897 |
| Baseline SERT binding | IL-15 | 66 | LMM: SERT BPP ~ immune marker + region + group + age + sex + BMI + GPAQ total + (1 \| subject) | 0.04 | [-0.171, 0.243] | 0.347 | 0.730 | 0.897 |
| Baseline SERT binding | IL-33 | 57 | LMM: SERT BPP ~ immune marker + region + group + age + sex + BMI + GPAQ total + (1 \| subject) | 0.03 | [-0.18, 0.229] | 0.243 | 0.809 | 0.897 |
| Baseline SERT binding | IL-22 | 65 | LMM: SERT BPP ~ immune marker + region + group + age + sex + BMI + GPAQ total + (1 \| subject) | 0.01 | [-0.16, 0.182] | 0.13 | 0.897 | 0.897 |
| Baseline subcortical volume | IL-33 | 56 | LMM: subcortical volume ~ immune marker + region + eTIV + group + age + sex + BMI + GPAQ total + (1 \| subject) | 0.16 | [-0.002, 0.322] | 1.989 | 0.052 | 0.378 |
| Baseline subcortical volume | IL-5 | 62 | LMM: subcortical volume ~ immune marker + region + eTIV + group + age + sex + BMI + GPAQ total + (1 \| subject) | -0.15 | [-0.327, 0.023] | -1.736 | 0.088 | 0.378 |
| Baseline subcortical volume | IL-13 | 58 | LMM: subcortical volume ~ immune marker + region + eTIV + group + age + sex + BMI + GPAQ total + (1 \| subject) | -0.14 | [-0.314, 0.034] | -1.611 | 0.113 | 0.378 |

| | | | | | | | | |
|---|---|---|---|---|---|---|---|---|
| Baseline subcortical volume | GFAP | 56 | LMM: subcortical volume ~ immune marker + region + eTIV + group + age + sex + BMI + GPAQ total + (1 \| subject) | 0.11 | [-0.075, 0.297] | 1.199 | 0.237 | 0.591 |
| Baseline subcortical volume | IL-8 | 62 | LMM: subcortical volume ~ immune marker + region + eTIV + group + age + sex + BMI + GPAQ total + (1 \| subject) | -0.10 | [-0.299, 0.107] | -0.95 | 0.346 | 0.691 |
| Baseline subcortical volume | IL-6 | 63 | LMM: subcortical volume ~ immune marker + region + eTIV + group + age + sex + BMI + GPAQ total + (1 \| subject) | -0.05 | [-0.211, 0.114] | -0.597 | 0.553 | 0.691 |
| Baseline subcortical volume | IL-22 | 63 | LMM: subcortical volume ~ immune marker + region + eTIV + group + age + sex + BMI + GPAQ total + (1 \| subject) | -0.04 | [-0.196, 0.107] | -0.587 | 0.560 | 0.691 |
| Baseline subcortical volume | IL-10 | 64 | LMM: subcortical volume ~ immune marker + region + eTIV + group + age + sex + BMI + GPAQ total + (1 \| subject) | -0.07 | [-0.32, 0.185] | -0.533 | 0.596 | 0.691 |
| Baseline subcortical volume | TNF | 64 | LMM: subcortical volume ~ immune marker + region + eTIV + group + age + sex + BMI + GPAQ total + (1 \| subject) | -0.05 | [-0.229, 0.138] | -0.496 | 0.622 | 0.691 |
| Baseline subcortical volume | IL-15 | 64 | LMM: subcortical volume ~ immune marker + region + eTIV + group + age + sex + BMI + GPAQ total + (1 \| subject) | -0.02 | [-0.193, 0.159] | -0.192 | 0.849 | 0.849 |
| Baseline cortical thickness | IL-6 | 63 | LMM: cortical thickness ~ immune marker + region + group + age + sex + BMI + GPAQ total + (1 \| subject) | -0.07 | [-0.255, 0.111] | -0.79 | 0.433 | 0.904 |
| Baseline cortical thickness | GFAP | 56 | LMM: cortical thickness ~ immune marker + region + group + age + sex + BMI + GPAQ total + (1 \| subject) | -0.06 | [-0.278, 0.152] | -0.588 | 0.559 | 0.904 |
| Baseline cortical thickness | IL-10 | 64 | LMM: cortical thickness ~ immune marker + region + group + age + sex + BMI + GPAQ total + (1 \| subject) | 0.08 | [-0.203, 0.369] | 0.582 | 0.563 | 0.904 |
| Baseline cortical thickness | IL-8 | 62 | LMM: cortical thickness ~ immune marker + region + group + age + sex + BMI + GPAQ total + (1 \| subject) | 0.06 | [-0.169, 0.291] | 0.534 | 0.596 | 0.904 |
| Baseline cortical thickness | IL-13 | 58 | LMM: cortical thickness ~ immune marker + region + group + age + sex + BMI + GPAQ total + (1 \| subject) | 0.05 | [-0.142, 0.245] | 0.53 | 0.599 | 0.904 |
| Baseline cortical thickness | IL-15 | 64 | LMM: cortical thickness ~ immune marker + region + group + age + sex + BMI + GPAQ total + (1 \| subject) | -0.05 | [-0.248, 0.15] | -0.495 | 0.623 | 0.904 |
| Baseline cortical thickness | TNF | 64 | LMM: cortical thickness ~ immune marker + region + group + age + sex + BMI + GPAQ total + (1 \| subject) | -0.05 | [-0.252, 0.154] | -0.48 | 0.633 | 0.904 |
| Baseline cortical thickness | IL-5 | 62 | LMM: cortical thickness ~ immune marker + region + group + age + sex + BMI + GPAQ total + (1 \| subject) | -0.02 | [-0.217, 0.185] | -0.158 | 0.875 | 0.973 |
| Baseline cortical thickness | IL-22 | 63 | LMM: cortical thickness ~ immune marker + region + group + age + sex + BMI + GPAQ total + (1 \| subject) | 0.01 | [-0.167, 0.177] | 0.055 | 0.956 | 0.973 |
| Baseline cortical thickness | IL-33 | 56 | LMM: cortical thickness ~ immune marker + region + group + age + sex + BMI + GPAQ total + (1 \| subject) | 0.00 | [-0.18, 0.186] | 0.034 | 0.973 | 0.973 |
| Hippocampal volume change with treatment | IL-15 | 24 | LMM: hippocampal volume ~ timepoint * immune marker + eTIV + age + sex + BMI + GPAQ total + (1 \| subject) | 103.81 | [19.177, 188.446] | 2.587 | 0.019 | 0.191 |

| | | | | | | | | |
|---|---|---|---|---|---|---|---|---|
| Hippocampal volume change with treatment | IL-13 | 21 | LMM: hippocampal volume ~ timepoint * immune marker + eTIV + age + sex + BMI + GPAQ total + (1 \| subject) | 66.47 | [-7.036, 139.966] | 1.939 | 0.073 | 0.364 |
| Hippocampal volume change with treatment | IL-22 | 23 | LMM: hippocampal volume ~ timepoint * immune marker + eTIV + age + sex + BMI + GPAQ total + (1 \| subject) | -49.05 | [-148.002, 49.912] | -1.046 | 0.310 | 0.820 |
| Hippocampal volume change with treatment | TNF | 24 | LMM: hippocampal volume ~ timepoint * immune marker + eTIV + age + sex + BMI + GPAQ total + (1 \| subject) | -32.53 | [-119.786, 54.733] | -0.786 | 0.442 | 0.820 |
| Hippocampal volume change with treatment | IL-5 | 24 | LMM: hippocampal volume ~ timepoint * immune marker + eTIV + age + sex + BMI + GPAQ total + (1 \| subject) | 25.28 | [-59.187, 109.75] | 0.631 | 0.536 | 0.820 |
| Hippocampal volume change with treatment | IL-6 | 24 | LMM: hippocampal volume ~ timepoint * immune marker + eTIV + age + sex + BMI + GPAQ total + (1 \| subject) | 17.29 | [-40.547, 75.132] | 0.631 | 0.537 | 0.820 |
| Hippocampal volume change with treatment | GFAP | 20 | LMM: hippocampal volume ~ timepoint * immune marker + eTIV + age + sex + BMI + GPAQ total + (1 \| subject) | 24.94 | [-110.353, 160.236] | 0.395 | 0.699 | 0.820 |
| Hippocampal volume change with treatment | IL-8 | 22 | LMM: hippocampal volume ~ timepoint * immune marker + eTIV + age + sex + BMI + GPAQ total + (1 \| subject) | -18.19 | [-153.793, 117.411] | -0.286 | 0.779 | 0.820 |
| Hippocampal volume change with treatment | IL-33 | 22 | LMM: hippocampal volume ~ timepoint * immune marker + eTIV + age + sex + BMI + GPAQ total + (1 \| subject) | 10.45 | [-81.148, 102.043] | 0.243 | 0.811 | 0.820 |
| Hippocampal volume change with treatment | IL-10 | 24 | LMM: hippocampal volume ~ timepoint * immune marker + eTIV + age + sex + BMI + GPAQ total + (1 \| subject) | -11.25 | [-114.166, 91.664] | -0.231 | 0.820 | 0.820 |

**Supplementary Table S4: Exploratory longitudinal associations of IL-13 and IL-15 with treatment outcomes measured using BDI and $HDRS_{17}$.** Linear mixed effects models (LMMs) tested the timepoint × immune marker. Both unadjusted models and models adjusted for age, sex, body mass index (BMI), and total score of Global Physical Activity Questionnaire (GPAQ) are presented. BDI, Beck Depression Inventory; $HDRS_{17}$, 17-item Hamilton Depression Rating Scale; IL, interleukin.

| β | SE | 95% CI low | 95% CI high | Statistic | p | n | Scale | Marker | Model |
|---|---|---|---|---|---|---|---|---|---|
| 3.97 | 1.46 | 0.98 | 6.95 | 2.72 | 0.011 | 33 | BDI | IL-13 | LMM: BDI ~ timepoint * immune marker + (1 \| subject) |
| 3.11 | 1.34 | 0.37 | 5.86 | 2.32 | 0.027 | 36 | BDI | IL-15 | LMM: BDI ~ timepoint * immune marker + (1 \| subject) |
| 3.20 | 1.88 | -0.72 | 7.11 | 1.70 | 0.104 | 22 | BDI | IL-13 | LMM: BDI ~ timepoint * immune marker + age + sex + BMI + GPAQ total + (1 \| subject) |
| 2.58 | 1.58 | -0.71 | 5.87 | 1.63 | 0.118 | 25 | BDI | IL-15 | LMM: BDI ~ timepoint * immune marker + age + sex + BMI + GPAQ total + (1 \| subject) |
| 4.04 | 1.14 | 1.71 | 6.37 | 3.53 | 0.001 | 33 | $HDRS_{17}$ | IL-13 | LMM: $HDRS_{17}$ ~ timepoint * immune marker + (1 \| subject) |
| 2.85 | 1.06 | 0.69 | 5.01 | 2.69 | 0.011 | 36 | $HDRS_{17}$ | IL-15 | LMM: $HDRS_{17}$ ~ timepoint * immune marker + (1 \| subject) |
| 3.88 | 1.43 | 0.92 | 6.85 | 2.72 | 0.013 | 22 | $HDRS_{17}$ | IL-13 | LMM: $HDRS_{17}$ ~ timepoint * immune marker + age + sex + BMI + GPAQ total + (1 \| subject) |
| 2.40 | 1.25 | -0.20 | 4.99 | 1.92 | 0.068 | 25 | $HDRS_{17}$ | IL-15 | LMM: $HDRS_{17}$ ~ timepoint * immune marker + age + sex + BMI + GPAQ total + (1 \| subject) |

**Supplementary Table S5: Omnibus tests of region × immune marker interactions in baseline neuroimaging outcomes.** Nested models specifying a common immune marker slope across regions were compared with models including a region × immune marker interaction to test whether associations varied by region. Results are presented for unadjusted models and models adjusted for age, sex, body mass index (BMI), and total score of Global Physical Activity Questionnaire (GPAQ). SERT, serotonin transporter; IL, interleukin; TNF, tumor necrosis factor.

| **Covariates** | **Domain** | **Marker** | **n** | **$\chi^2$** | **df** | **p** |
|---|---|---|---|---|---|---|
| none | Baseline SERT binding | IL-5 | 75 | 2.87 | 5 | 0.720 |
| none | Baseline SERT binding | TNF | 78 | 2.28 | 5 | 0.809 |
| none | Baseline subcortical volume | IL-5 | 75 | 2.72 | 3 | 0.437 |
| none | Baseline subcortical volume | IL-33 | 69 | 1.90 | 3 | 0.592 |
| none | Baseline cortical thickness | IL-15 | 78 | 0.40 | 3 | 0.939 |
| age + sex + BMI + GPAQ total | Baseline SERT binding | IL-5 | 64 | 2.39 | 5 | 0.794 |
| age + sex + BMI + GPAQ total | Baseline SERT binding | TNF | 66 | 1.30 | 5 | 0.935 |
| age + sex + BMI + GPAQ total | Baseline subcortical volume | IL-5 | 62 | 4.82 | 3 | 0.185 |
| age + sex + BMI + GPAQ total | Baseline subcortical volume | IL-33 | 56 | 3.85 | 3 | 0.278 |
| age + sex + BMI + GPAQ total | Baseline cortical thickness | IL-15 | 64 | 0.83 | 3 | 0.842 |

**Supplementary Table S6: Leave-one-region-out sensitivity analyses of immune marker associations with baseline neuroimaging outcomes.** Each primary model was refitted after sequentially excluding one brain region to assess whether the estimated immune marker association was driven by a particular region. SERT, serotonin transporter; IL, interleukin; TNF, tumor necrosis factor.

| | | Primary model statistics | | | | | | Leave-one-region-out effects | | | | | | |
|---|---|---|---|---|---|---|---|---|---|---|---|---|---|---|
| **Domain** | **Marker** | **β** | **95%CI low** | **95%CI high** | **p** | **n** | **n (regions)** | **Min. β** | **Max. β** | **Max. absolute change from primary** | **Regions producing maximum change** | **Directions agree with primary** | **Min. p** | **Max. p** |
| Baseline cortical thickness | IL-15 | -0.23 | -0.39 | -0.06 | 0.007 | 78 | 4 | -0.25 | -0.22 | 0.03 | Caudal anterior cingulate cortex | all | 0.005 | 0.017 |
| Baseline subcortical volume | IL-33 | 0.22 | 0.07 | 0.38 | 0.005 | 69 | 4 | 0.21 | 0.25 | 0.02 | Caudate | all | 0.004 | 0.010 |
| Baseline subcortical volume | IL-5 | -0.14 | -0.31 | 0.02 | 0.085 | 75 | 4 | -0.16 | -0.12 | 0.02 | Hippocampus | all | 0.069 | 0.167 |
| Baseline SERT binding | TNF | -0.27 | -0.48 | -0.07 | 0.010 | 78 | 6 | -0.29 | -0.26 | 0.02 | Anterior cingulate cortex | all | 0.008 | 0.014 |

**Supplementary Table S7: Leave-one-participant-out sensitivity analyses of immune-marker associations.** Each significant primary model was repeatedly refitted after excluding one participant at a time to assess the influence of individual observations. p, nominal p; $p_{FDR,domain}$, FDR-adjusted within the corresponding domain; $p_{FDR,global}$, FDR-adjusted across all primary analyses. GFAP, glial fibrillary acidic protein; IL, interleukin; SERT, serotonin transporter; TNF, tumor necrosis factor.

| | | | **Primary model statistics** | | | | | **Leave one participant out effects** | | | | | | |
|---|---|---|---|---|---|---|---|---|---|---|---|---|---|---|
| **Domain** | **Covariates** | **Marker** | **β** | **p** | **$p_{FDR,domain}$** | **$p_{FDR,global}$** | **n** | **n refits** | **n converged refits** | **min. β** | **max. β** | **max. absolute change** | **sign reversal** | **proportion nominally significant** |
| Diagnosis | none | IL-6 | 0.55 | 0.012 | 0.054 | 0.105 | 83 | 83 | 83 | 0.43 | 0.61 | 0.12 | FALSE | 1.00 |
| Diagnosis | none | TNF | 0.52 | 0.016 | 0.054 | 0.123 | 84 | 84 | 84 | 0.46 | 0.59 | 0.07 | FALSE | 1.00 |
| Diagnosis | none | IL-8 | 0.49 | 0.029 | 0.072 | 0.173 | 80 | 80 | 80 | 0.43 | 0.59 | 0.10 | FALSE | 0.99 |
| Diagnosis | none | IL-33 | -0.48 | 0.041 | 0.081 | 0.203 | 74 | 74 | 74 | -0.53 | -0.41 | 0.07 | FALSE | 0.74 |
| Diagnosis | none | GFAP | 0.66 | 0.004 | 0.042 | 0.100 | 75 | 75 | 75 | 0.59 | 0.73 | 0.07 | FALSE | 1.00 |
| Baseline SERT binding | none | TNF | -0.27 | 0.010 | 0.105 | 0.105 | 78 | 78 | 78 | -0.34 | -0.20 | 0.07 | FALSE | 0.99 |
| Antidepressant response | none | IL-13 | 6.22 | 0.000 | 0.002 | 0.011 | 33 | 33 | 33 | 5.27 | 7.29 | 1.07 | FALSE | 1.00 |
| Antidepressant response | none | IL-15 | 3.29 | 0.034 | 0.169 | 0.184 | 36 | 36 | 36 | 2.22 | 6.53 | 3.25 | FALSE | 0.89 |
| Baseline subcortical volume | none | IL-33 | 0.22 | 0.005 | 0.050 | 0.100 | 69 | 69 | 69 | 0.20 | 0.26 | 0.04 | FALSE | 1.00 |
| Baseline cortical thickness | none | IL-15 | -0.23 | 0.007 | 0.071 | 0.105 | 78 | 78 | 78 | -0.27 | -0.20 | 0.04 | FALSE | 1.00 |
| Hippocampal volume change with treatment | none | IL-13 | 64.44 | 0.023 | 0.117 | 0.156 | 31 | 31 | 31 | 57.65 | 76.33 | 11.89 | FALSE | 1.00 |
| Hippocampal volume change with treatment | none | IL-15 | 81.64 | 0.012 | 0.117 | 0.105 | 34 | 34 | 34 | 73.67 | 99.25 | 17.62 | FALSE | 1.00 |

**Supplementary Table S8: Exploratory associations of IL-17A detectability with clinical, serotonergic, and structural neuroimaging measures.** IL-17A concentrations were categorized as detectable or non-detectable, and analyses were performed using unadjusted models and models adjusted for age, sex, body mass index (BMI), and total score of Global Physical Activity Questionnaire (GPAQ). Logistic regression was used for diagnosis, with the odds ratio (OR) obtained by exponentiating β. Longitudinal models tested the timepoint × IL-17A detectability interaction. MADRS, Montgomery-Åsberg Depression Rating Scale; SERT, serotonin transporter; $BP_P$, binding potential relative to plasma; eTIV, estimated total intracranial volume.

| Domain | n | n (detected) | n (not detected) | Model | β | SE | 95% CI low | 95% CI high | statistic | p | OR | OR CI low | OR CI high |
|---|---|---|---|---|---|---|---|---|---|---|---|---|---|
| Diagnosis | 84 | 65 | 19 | Logistic regression: IL-17A detectability ~ diagnosis | 1.29 | 0.61 | 0.16 | 2.63 | 2.10 | 0.036 | 3.64 | 1.17 | 13.82 |
| Antidepressant response | 36 | 32 | 4 | LMM: MADRS ~ timepoint * IL-17A detectability + (1 \| subject) | -8.89 | 5.61 | -20.21 | 2.43 | -1.58 | 0.121 | - | - | - |
| SERT binding | 78 | 61 | 17 | LMM: SERT $BP_P$ ~ IL-17A detectability + region + group + (1 \| subject) | -0.21 | 0.25 | -0.71 | 0.28 | -0.86 | 0.391 | - | - | - |
| Baseline subcortical volume | 78 | 63 | 15 | LMM: subcortical volume ~ IL-17A detectability + region + eTIV + group + (1 \| subject) | 0.06 | 0.20 | -0.33 | 0.45 | 0.32 | 0.749 | - | - | - |
| Baseline cortical thickness | 78 | 63 | 15 | LMM: cortical thickness ~ IL-17A detectability + region + group + (1 \| subject) | 0.43 | 0.22 | 0.00 | 0.87 | 1.99 | 0.050 | - | - | - |
| Hippocampal volume change with treatment | 34 | 31 | 3 | LMM: hippocampal volume ~ timepoint * IL-17A detectability + eTIV + (1 \| subject) | -79.02 | 103.75 | -292.65 | 134.60 | -0.76 | 0.453 | - | - | - |
| Diagnosis | 69 | 51 | 18 | Logistic regression: IL-17A detectability ~ diagnosis + age + sex + BMI + GPAQ total | 1.13 | 0.70 | -0.16 | 2.64 | 1.61 | 0.107 | 3.09 | 0.85 | 14.06 |
| Antidepressant response | 25 | 21 | 4 | LMM: MADRS ~ timepoint * IL-17A detectability + age + sex + BMI + GPAQ total + (1 \| subject) | -8.87 | 6.32 | -21.83 | 4.10 | -1.40 | 0.172 | - | - | - |
| SERT binding | 66 | 50 | 16 | LMM: SERT $BP_P$ ~ IL-17A detectability + region + group + age + sex + BMI + GPAQ total + (1 \| subject) | -0.06 | 0.22 | -0.49 | 0.37 | -0.28 | 0.781 | - | - | - |

| | | | | | | | | | | | | | |
|---|---|---|---|---|---|---|---|---|---|---|---|---|---|
| Baseline subcortical volume | 64 | 50 | 14 | LMM: subcortical volume ~ IL-17A detectability + region + eTIV + group + age + sex + BMI + GPAQ total + (1 \| subject) | -0.07 | 0.20 | -0.46 | 0.32 | -0.36 | 0.718 | - | - | - |
| Baseline cortical thickness | 64 | 50 | 14 | LMM: cortical thickness ~ IL-17A detectability + region + group + age + sex + BMI + GPAQ total + (1 \| subject) | 0.50 | 0.21 | 0.09 | 0.92 | 2.41 | 0.019 | - | - | - |
| Hippocampal volume change with treatment | 24 | 21 | 3 | LMM: hippocampal volume ~ timepoint * IL-17A detectability + eTIV + age + sex + BMI + GPAQ total + (1 \| subject) | -76.90 | 114.99 | -319.43 | 165.64 | -0.67 | 0.513 | - | - | - |

**Supplementary Table S10: Exploratory associations of routine peripheral inflammatory markers with clinical, serotonergic, and structural neuroimaging measures.** Associations of C-reactive protein (CRP), fibrinogen, leukocyte subsets, and platelet counts with diagnosis, antidepressant response, baseline serotonin transporter (SERT) binding, baseline subcortical volume, baseline cortical thickness, and treatment-related hippocampal volume change were examined using unadjusted models and models adjusted for age, sex, body mass index (BMI), and total score of Global Physical Activity Questionnaire (GPAQ). Longitudinal models tested the timepoint × routine marker interaction. MADRS, Montgomery-Åsberg Depression Rating Scale; $BP_P$, binding potential relative to plasma; eTIV, estimated total intracranial volume.

| Domain | Marker | Covariates | n | Model | β | SE | 95% CI low | 95%CI high | statistic | p |
|---|---|---|---|---|---|---|---|---|---|---|
| Diagnosis | CRP | none | 80 | routine marker ~ diagnosis | 0.08 | 0.23 | -0.37 | 0.53 | 0.35 | 0.730 |
| Diagnosis | Fibrinogen | none | 74 | routine marker ~ diagnosis | 0.32 | 0.23 | -0.14 | 0.78 | 1.37 | 0.174 |
| Diagnosis | Leukocytes | none | 83 | routine marker ~ diagnosis | 0.62 | 0.21 | 0.20 | 1.04 | 2.92 | 0.005 |
| Diagnosis | Neutrophils | none | 77 | routine marker ~ diagnosis | 0.35 | 0.23 | -0.11 | 0.80 | 1.52 | 0.133 |
| Diagnosis | Lymphocytes | none | 77 | routine marker ~ diagnosis | 0.64 | 0.22 | 0.20 | 1.07 | 2.90 | 0.005 |
| Diagnosis | Monocytes | none | 77 | routine marker ~ diagnosis | 0.35 | 0.23 | -0.10 | 0.81 | 1.56 | 0.123 |
| Diagnosis | Eosinophils | none | 77 | routine marker ~ diagnosis | 0.39 | 0.23 | -0.06 | 0.84 | 1.73 | 0.088 |
| Diagnosis | Platelets | none | 83 | routine marker ~ diagnosis | 0.42 | 0.22 | -0.01 | 0.86 | 1.93 | 0.058 |
| Antidepressant response | CRP | none | 34 | LMM: MADRS ~ timepoint * routine marker + (1 \| subject) | 1.45 | 1.85 | -2.29 | 5.18 | 0.78 | 0.438 |
| Antidepressant response | Fibrinogen | none | 35 | LMM: MADRS ~ timepoint * routine marker + (1 \| subject) | -0.39 | 1.65 | -3.75 | 2.97 | -0.24 | 0.815 |
| Antidepressant response | Leukocytes | none | 35 | LMM: MADRS ~ timepoint * routine marker + (1 \| subject) | 2.08 | 1.64 | -1.28 | 5.44 | 1.27 | 0.215 |
| Antidepressant response | Neutrophils | none | 34 | LMM: MADRS ~ timepoint * routine marker + (1 \| subject) | 2.14 | 1.70 | -1.33 | 5.61 | 1.26 | 0.217 |

| | | | | | | | | | | |
|---|---|---|---|---|---|---|---|---|---|---|
| Antidepressant response | Lymphocytes | none | 34 | LMM: MADRS ~ timepoint * routine marker + (1 \| subject) | -0.14 | 1.65 | -3.52 | 3.24 | -0.08 | 0.934 |
| Antidepressant response | Monocytes | none | 34 | LMM: MADRS ~ timepoint * routine marker + (1 \| subject) | 2.30 | 1.59 | -0.96 | 5.55 | 1.44 | 0.160 |
| Antidepressant response | Eosinophils | none | 34 | LMM: MADRS ~ timepoint * routine marker + (1 \| subject) | 1.24 | 1.80 | -2.43 | 4.91 | 0.69 | 0.497 |
| Antidepressant response | Platelets | none | 35 | LMM: MADRS ~ timepoint * routine marker + (1 \| subject) | 1.70 | 1.61 | -1.58 | 4.98 | 1.06 | 0.298 |
| Baseline SERT binding | CRP | none | 74 | LMM: SERT $BP_P$ ~ routine marker + region + group + (1 \| subject) | 0.17 | 0.11 | -0.06 | 0.39 | 1.49 | 0.141 |
| Baseline SERT binding | Fibrinogen | none | 68 | LMM: SERT $BP_P$ ~ routine marker + region + group + (1 \| subject) | 0.11 | 0.11 | -0.11 | 0.34 | 1.02 | 0.312 |
| Baseline SERT binding | Leukocytes | none | 77 | LMM: SERT $BP_P$ ~ routine marker + region + group + (1 \| subject) | 0.09 | 0.10 | -0.12 | 0.30 | 0.89 | 0.378 |
| Baseline SERT binding | Neutrophils | none | 72 | LMM: SERT $BP_P$ ~ routine marker + region + group + (1 \| subject) | 0.06 | 0.10 | -0.15 | 0.27 | 0.57 | 0.567 |
| Baseline SERT binding | Lymphocytes | none | 72 | LMM: SERT $BP_P$ ~ routine marker + region + group + (1 \| subject) | 0.13 | 0.11 | -0.09 | 0.34 | 1.19 | 0.236 |
| Baseline SERT binding | Monocytes | none | 72 | LMM: SERT $BP_P$ ~ routine marker + region + group + (1 \| subject) | -0.07 | 0.10 | -0.27 | 0.14 | -0.64 | 0.523 |
| Baseline SERT binding | Eosinophils | none | 72 | LMM: SERT $BP_P$ ~ routine marker + region + group + (1 \| subject) | 0.04 | 0.11 | -0.18 | 0.26 | 0.36 | 0.719 |
| Baseline SERT binding | Platelets | none | 77 | LMM: SERT $BP_P$ ~ routine marker + region + group + (1 \| subject) | 0.11 | 0.10 | -0.09 | 0.31 | 1.11 | 0.271 |
| Baseline subcortical volume | CRP | none | 74 | LMM: subcortical volume ~ routine marker + region + group + eTIV + (1 \| subject) | -0.01 | 0.08 | -0.17 | 0.15 | -0.13 | 0.895 |
| Baseline subcortical volume | Fibrinogen | none | 68 | LMM: subcortical volume ~ routine marker + region + group + eTIV + (1 \| subject) | -0.03 | 0.09 | -0.20 | 0.14 | -0.32 | 0.752 |
| Baseline subcortical volume | Leukocytes | none | 77 | LMM: subcortical volume ~ routine marker + region + group + eTIV + (1 \| subject) | 0.06 | 0.08 | -0.10 | 0.22 | 0.80 | 0.427 |
| Baseline subcortical volume | Neutrophils | none | 72 | LMM: subcortical volume ~ routine marker + region + group + eTIV + (1 \| subject) | 0.00 | 0.08 | -0.16 | 0.15 | -0.05 | 0.962 |

| | | | | | | | | | | |
|---|---|---|---|---|---|---|---|---|---|---|
| Baseline subcortical volume | Lymphocytes | none | 72 | LMM: subcortical volume ~ routine marker + region + group + eTIV + (1 \| subject) | 0.06 | 0.08 | -0.10 | 0.23 | 0.77 | 0.443 |
| Baseline subcortical volume | Monocytes | none | 72 | LMM: subcortical volume ~ routine marker + region + group + eTIV + (1 \| subject) | 0.12 | 0.08 | -0.03 | 0.28 | 1.60 | 0.113 |
| Baseline subcortical volume | Eosinophils | none | 72 | LMM: subcortical volume ~ routine marker + region + group + eTIV + (1 \| subject) | 0.04 | 0.08 | -0.12 | 0.19 | 0.48 | 0.630 |
| Baseline subcortical volume | Platelets | none | 77 | LMM: subcortical volume ~ routine marker + region + group + eTIV + (1 \| subject) | 0.04 | 0.08 | -0.11 | 0.19 | 0.58 | 0.562 |
| Baseline cortical thickness | CRP | none | 74 | LMM: cortical thickness ~ routine marker + region + group + (1 \| subject) | 0.11 | 0.09 | -0.08 | 0.29 | 1.13 | 0.262 |
| Baseline cortical thickness | Fibrinogen | none | 68 | LMM: cortical thickness ~ routine marker + region + group + (1 \| subject) | 0.03 | 0.09 | -0.16 | 0.22 | 0.31 | 0.761 |
| Baseline cortical thickness | Leukocytes | none | 77 | LMM: cortical thickness ~ routine marker + region + group + (1 \| subject) | -0.11 | 0.09 | -0.30 | 0.07 | -1.25 | 0.217 |
| Baseline cortical thickness | Neutrophils | none | 72 | LMM: cortical thickness ~ routine marker + region + group + (1 \| subject) | -0.15 | 0.09 | -0.33 | 0.03 | -1.65 | 0.104 |
| Baseline cortical thickness | Lymphocytes | none | 72 | LMM: cortical thickness ~ routine marker + region + group + (1 \| subject) | 0.14 | 0.09 | -0.05 | 0.32 | 1.45 | 0.151 |
| Baseline cortical thickness | Monocytes | none | 72 | LMM: cortical thickness ~ routine marker + region + group + (1 \| subject) | -0.02 | 0.09 | -0.21 | 0.16 | -0.27 | 0.789 |
| Baseline cortical thickness | Eosinophils | none | 72 | LMM: cortical thickness ~ routine marker + region + group + (1 \| subject) | 0.09 | 0.09 | -0.09 | 0.27 | 1.03 | 0.305 |
| Baseline cortical thickness | Platelets | none | 77 | LMM: cortical thickness ~ routine marker + region + group + (1 \| subject) | -0.09 | 0.09 | -0.27 | 0.08 | -1.08 | 0.282 |
| Hippocampal volume change with treatment | CRP | none | 32 | LMM: hippocampal volume ~ timepoint * routine marker + eTIV + (1 \| subject) | 38.17 | 37.83 | -40.07 | 116.41 | 1.01 | 0.323 |
| Hippocampal volume change with treatment | Fibrinogen | none | 33 | LMM: hippocampal volume ~ timepoint * routine marker + eTIV + (1 \| subject) | 50.61 | 31.72 | -14.84 | 116.06 | 1.60 | 0.124 |

| | | | | | | | | | | |
|---|---|---|---|---|---|---|---|---|---|---|
| Hippocampal volume change with treatment | Leukocytes | none | 33 | LMM: hippocampal volume ~ timepoint * routine marker + eTIV + (1 \| subject) | 19.66 | 29.35 | -40.90 | 80.23 | 0.67 | 0.509 |
| Hippocampal volume change with treatment | Neutrophils | none | 33 | LMM: hippocampal volume ~ timepoint * routine marker + eTIV + (1 \| subject) | 31.80 | 28.67 | -27.37 | 90.97 | 1.11 | 0.278 |
| Hippocampal volume change with treatment | Lymphocytes | none | 33 | LMM: hippocampal volume ~ timepoint * routine marker + eTIV + (1 \| subject) | -32.21 | 30.18 | -94.49 | 30.07 | -1.07 | 0.296 |
| Hippocampal volume change with treatment | Monocytes | none | 33 | LMM: hippocampal volume ~ timepoint * routine marker + eTIV + (1 \| subject) | 29.93 | 28.38 | -28.63 | 88.49 | 1.05 | 0.302 |
| Hippocampal volume change with treatment | Eosinophils | none | 33 | LMM: hippocampal volume ~ timepoint * routine marker + eTIV + (1 \| subject) | 22.01 | 33.63 | -47.38 | 91.40 | 0.65 | 0.519 |
| Hippocampal volume change with treatment | Platelets | none | 33 | LMM: hippocampal volume ~ timepoint * routine marker + eTIV + (1 \| subject) | 36.86 | 34.06 | -33.41 | 107.13 | 1.08 | 0.290 |
| Diagnosis | CRP | age + sex + BMI + GPAQ total | 65 | routine marker ~ group + age + sex + BMI + GPAQ total | 0.01 | 0.24 | -0.48 | 0.50 | 0.04 | 0.967 |
| Diagnosis | Fibrinogen | age + sex + BMI + GPAQ total | 61 | routine marker ~ group + age + sex + BMI + GPAQ total | 0.50 | 0.28 | -0.05 | 1.05 | 1.82 | 0.075 |
| Diagnosis | Leukocytes | age + sex + BMI + GPAQ total | 68 | routine marker ~ group + age + sex + BMI + GPAQ total | 0.73 | 0.23 | 0.26 | 1.20 | 3.13 | 0.003 |
| Diagnosis | Neutrophils | age + sex + BMI + GPAQ total | 63 | routine marker ~ group + age + sex + BMI + GPAQ total | 0.41 | 0.26 | -0.11 | 0.93 | 1.59 | 0.118 |
| Diagnosis | Lymphocytes | age + sex + BMI + GPAQ total | 63 | routine marker ~ group + age + sex + BMI + GPAQ total | 0.55 | 0.26 | 0.02 | 1.08 | 2.09 | 0.041 |
| Diagnosis | Monocytes | age + sex + BMI + GPAQ total | 63 | routine marker ~ group + age + sex + BMI + GPAQ total | 0.47 | 0.25 | -0.04 | 0.97 | 1.85 | 0.069 |
| Diagnosis | Eosinophils | age + sex + BMI + GPAQ total | 63 | routine marker ~ group + age + sex + BMI + GPAQ total | 0.49 | 0.29 | -0.09 | 1.06 | 1.70 | 0.094 |
| Diagnosis | Platelets | age + sex + BMI + GPAQ total | 68 | routine marker ~ group + age + sex + BMI + GPAQ total | 0.56 | 0.22 | 0.11 | 1.01 | 2.51 | 0.015 |
| Antidepressant response | CRP | age + sex + BMI + GPAQ total | 23 | LMM: MADRS ~ timepoint * routine marker + age + sex + BMI + GPAQ total + (1 \| subject) | 0.33 | 2.22 | -4.30 | 4.95 | 0.15 | 0.885 |

| | | | | | | | | | | |
|---|---|---|---|---|---|---|---|---|---|---|
| Antidepressant response | Fibrinogen | age + sex + BMI + GPAQ total | 24 | LMM: MADRS ~ timepoint * routine marker + age + sex + BMI + GPAQ total + (1 \| subject) | -1.24 | 1.99 | -5.39 | 2.91 | -0.62 | 0.541 |
| Antidepressant response | Leukocytes | age + sex + BMI + GPAQ total | 24 | LMM: MADRS ~ timepoint * routine marker + age + sex + BMI + GPAQ total + (1 \| subject) | 1.43 | 2.60 | -3.98 | 6.85 | 0.55 | 0.588 |
| Antidepressant response | Neutrophils | age + sex + BMI + GPAQ total | 24 | LMM: MADRS ~ timepoint * routine marker + age + sex + BMI + GPAQ total + (1 \| subject) | 2.08 | 2.38 | -2.89 | 7.05 | 0.87 | 0.393 |
| Antidepressant response | Lymphocytes | age + sex + BMI + GPAQ total | 24 | LMM: MADRS ~ timepoint * routine marker + age + sex + BMI + GPAQ total + (1 \| subject) | -2.01 | 2.03 | -6.24 | 2.23 | -0.99 | 0.335 |
| Antidepressant response | Monocytes | age + sex + BMI + GPAQ total | 24 | LMM: MADRS ~ timepoint * routine marker + age + sex + BMI + GPAQ total + (1 \| subject) | 1.31 | 2.52 | -3.94 | 6.55 | 0.52 | 0.610 |
| Antidepressant response | Eosinophils | age + sex + BMI + GPAQ total | 24 | LMM: MADRS ~ timepoint * routine marker + age + sex + BMI + GPAQ total + (1 \| subject) | 0.80 | 2.18 | -3.71 | 5.32 | 0.37 | 0.716 |
| Antidepressant response | Platelets | age + sex + BMI + GPAQ total | 24 | LMM: MADRS ~ timepoint * routine marker + age + sex + BMI + GPAQ total + (1 \| subject) | 1.07 | 1.92 | -2.94 | 5.08 | 0.56 | 0.584 |
| Baseline SERT binding | CRP | age + sex + BMI + GPAQ total | 62 | LMM: SERT $BP_P$ ~ routine marker + region + group + age + sex + BMI + GPAQ total + (1 \| subject) | 0.14 | 0.10 | -0.07 | 0.35 | 1.33 | 0.190 |
| Baseline SERT binding | Fibrinogen | age + sex + BMI + GPAQ total | 58 | LMM: SERT $BP_P$ ~ routine marker + region + group + age + sex + BMI + GPAQ total + (1 \| subject) | 0.13 | 0.10 | -0.07 | 0.33 | 1.32 | 0.192 |
| Baseline SERT binding | Leukocytes | age + sex + BMI + GPAQ total | 65 | LMM: SERT $BP_P$ ~ routine marker + region + group + age + sex + BMI + GPAQ total + (1 \| subject) | 0.05 | 0.11 | -0.17 | 0.26 | 0.43 | 0.669 |
| Baseline SERT binding | Neutrophils | age + sex + BMI + GPAQ total | 60 | LMM: SERT $BP_P$ ~ routine marker + region + group + age + sex + BMI + GPAQ total + (1 \| subject) | 0.07 | 0.10 | -0.13 | 0.28 | 0.71 | 0.483 |
| Baseline SERT binding | Lymphocytes | age + sex + BMI + GPAQ total | 60 | LMM: SERT $BP_P$ ~ routine marker + region + group + age + sex + BMI + GPAQ total + (1 \| subject) | -0.05 | 0.10 | -0.25 | 0.15 | -0.51 | 0.615 |
| Baseline SERT binding | Monocytes | age + sex + BMI + GPAQ total | 60 | LMM: SERT $BP_P$ ~ routine marker + region + group + age + sex + BMI + GPAQ total + (1 \| subject) | -0.07 | 0.10 | -0.27 | 0.14 | -0.65 | 0.521 |
| Baseline SERT binding | Eosinophils | age + sex + BMI + GPAQ total | 60 | LMM: SERT $BP_P$ ~ routine marker + region + group + age + sex + BMI + GPAQ total + (1 \| subject) | 0.08 | 0.09 | -0.11 | 0.26 | 0.83 | 0.413 |
| Baseline SERT binding | Platelets | age + sex + BMI + GPAQ total | 65 | LMM: SERT $BP_P$ ~ routine marker + region + group + age + sex + BMI + GPAQ total + (1 \| subject) | 0.10 | 0.11 | -0.11 | 0.32 | 0.96 | 0.343 |
| Baseline subcortical volume | CRP | age + sex + BMI + GPAQ total | 60 | LMM: subcortical volume ~ routine marker + region + group + eTIV + age + sex + BMI + GPAQ total + (1 \| subject) | -0.12 | 0.09 | -0.29 | 0.06 | -1.33 | 0.188 |
| Baseline subcortical volume | Fibrinogen | age + sex + BMI + GPAQ total | 56 | LMM: subcortical volume ~ routine marker + region + group + eTIV + age + sex + BMI + GPAQ total + (1 \| subject) | -0.02 | 0.08 | -0.19 | 0.15 | -0.26 | 0.798 |

| | | | | | | | | | | |
|---|---|---|---|---|---|---|---|---|---|---|
| Baseline subcortical volume | Leukocytes | age + sex + BMI + GPAQ total | 63 | LMM: subcortical volume ~ routine marker + region + group + eTIV + age + sex + BMI + GPAQ total + (1 \| subject) | 0.06 | 0.09 | -0.13 | 0.24 | 0.62 | 0.535 |
| Baseline subcortical volume | Neutrophils | age + sex + BMI + GPAQ total | 58 | LMM: subcortical volume ~ routine marker + region + group + eTIV + age + sex + BMI + GPAQ total + (1 \| subject) | -0.02 | 0.09 | -0.20 | 0.17 | -0.21 | 0.836 |
| Baseline subcortical volume | Lymphocytes | age + sex + BMI + GPAQ total | 58 | LMM: subcortical volume ~ routine marker + region + group + eTIV + age + sex + BMI + GPAQ total + (1 \| subject) | -0.05 | 0.09 | -0.22 | 0.12 | -0.57 | 0.572 |
| Baseline subcortical volume | Monocytes | age + sex + BMI + GPAQ total | 58 | LMM: subcortical volume ~ routine marker + region + group + eTIV + age + sex + BMI + GPAQ total + (1 \| subject) | 0.15 | 0.09 | -0.03 | 0.32 | 1.67 | 0.101 |
| Baseline subcortical volume | Eosinophils | age + sex + BMI + GPAQ total | 58 | LMM: subcortical volume ~ routine marker + region + group + eTIV + age + sex + BMI + GPAQ total + (1 \| subject) | 0.00 | 0.08 | -0.15 | 0.16 | 0.05 | 0.961 |
| Baseline subcortical volume | Platelets | age + sex + BMI + GPAQ total | 63 | LMM: subcortical volume ~ routine marker + region + group + eTIV + age + sex + BMI + GPAQ total + (1 \| subject) | 0.09 | 0.09 | -0.09 | 0.28 | 1.05 | 0.297 |
| Baseline cortical thickness | CRP | age + sex + BMI + GPAQ total | 60 | LMM: cortical thickness ~ routine marker + region + group + age + sex + BMI + GPAQ total + (1 \| subject) | 0.04 | 0.10 | -0.16 | 0.24 | 0.40 | 0.688 |
| Baseline cortical thickness | Fibrinogen | age + sex + BMI + GPAQ total | 56 | LMM: cortical thickness ~ routine marker + region + group + age + sex + BMI + GPAQ total + (1 \| subject) | 0.04 | 0.09 | -0.15 | 0.23 | 0.40 | 0.691 |
| Baseline cortical thickness | Leukocytes | age + sex + BMI + GPAQ total | 63 | LMM: cortical thickness ~ routine marker + region + group + age + sex + BMI + GPAQ total + (1 \| subject) | -0.12 | 0.10 | -0.33 | 0.08 | -1.22 | 0.227 |
| Baseline cortical thickness | Neutrophils | age + sex + BMI + GPAQ total | 58 | LMM: cortical thickness ~ routine marker + region + group + age + sex + BMI + GPAQ total + (1 \| subject) | -0.12 | 0.10 | -0.32 | 0.08 | -1.18 | 0.244 |
| Baseline cortical thickness | Lymphocytes | age + sex + BMI + GPAQ total | 58 | LMM: cortical thickness ~ routine marker + region + group + age + sex + BMI + GPAQ total + (1 \| subject) | 0.00 | 0.10 | -0.19 | 0.20 | 0.04 | 0.972 |
| Baseline cortical thickness | Monocytes | age + sex + BMI + GPAQ total | 58 | LMM: cortical thickness ~ routine marker + region + group + age + sex + BMI + GPAQ total + (1 \| subject) | -0.06 | 0.10 | -0.26 | 0.15 | -0.55 | 0.585 |
| Baseline cortical thickness | Eosinophils | age + sex + BMI + GPAQ total | 58 | LMM: cortical thickness ~ routine marker + region + group + age + sex + BMI + GPAQ total + (1 \| subject) | 0.07 | 0.09 | -0.11 | 0.24 | 0.79 | 0.434 |
| Baseline cortical thickness | Platelets | age + sex + BMI + GPAQ total | 63 | LMM: cortical thickness ~ routine marker + region + group + age + sex + BMI + GPAQ total + (1 \| subject) | -0.18 | 0.10 | -0.38 | 0.02 | -1.76 | 0.084 |

| | | | | | | | | | | |
|---|---|---|---|---|---|---|---|---|---|---|
| Hippocampal volume change with treatment | CRP | age + sex + BMI + GPAQ total | 22 | LMM: hippocampal volume ~ timepoint * routine marker + eTIV + age + sex + BMI + GPAQ total + (1 \| subject) | 33.05 | 44.45 | -61.65 | 127.76 | 0.74 | 0.469 |
| Hippocampal volume change with treatment | Fibrinogen | age + sex + BMI + GPAQ total | 23 | LMM: hippocampal volume ~ timepoint * routine marker + eTIV + age + sex + BMI + GPAQ total + (1 \| subject) | 35.93 | 39.78 | -48.36 | 120.21 | 0.90 | 0.380 |
| Hippocampal volume change with treatment | Leukocytes | age + sex + BMI + GPAQ total | 23 | LMM: hippocampal volume ~ timepoint * routine marker + eTIV + age + sex + BMI + GPAQ total + (1 \| subject) | -13.99 | 52.38 | -125.00 | 97.01 | -0.27 | 0.793 |
| Hippocampal volume change with treatment | Neutrophils | age + sex + BMI + GPAQ total | 23 | LMM: hippocampal volume ~ timepoint * routine marker + eTIV + age + sex + BMI + GPAQ total + (1 \| subject) | 20.89 | 43.60 | -71.52 | 113.30 | 0.48 | 0.638 |
| Hippocampal volume change with treatment | Lymphocytes | age + sex + BMI + GPAQ total | 23 | LMM: hippocampal volume ~ timepoint * routine marker + eTIV + age + sex + BMI + GPAQ total + (1 \| subject) | -74.83 | 39.31 | -158.15 | 8.49 | -1.90 | 0.075 |
| Hippocampal volume change with treatment | Monocytes | age + sex + BMI + GPAQ total | 23 | LMM: hippocampal volume ~ timepoint * routine marker + eTIV + age + sex + BMI + GPAQ total + (1 \| subject) | -2.43 | 51.26 | -111.05 | 106.19 | -0.05 | 0.963 |
| Hippocampal volume change with treatment | Eosinophils | age + sex + BMI + GPAQ total | 23 | LMM: hippocampal volume ~ timepoint * routine marker + eTIV + age + sex + BMI + GPAQ total + (1 \| subject) | -7.68 | 42.55 | -97.84 | 82.48 | -0.18 | 0.859 |
| Hippocampal volume change with treatment | Platelets | age + sex + BMI + GPAQ total | 23 | LMM: hippocampal volume ~ timepoint * routine marker + eTIV + age + sex + BMI + GPAQ total + (1 \| subject) | 23.94 | 46.62 | -74.83 | 122.71 | 0.51 | 0.615 |

### Supplementary Figures

**Supplementary Figure S1: CONSORT flow diagram.** MDD, major depressive disorder; MRI, magnetic resonance imaging; PET, positron emission tomography.

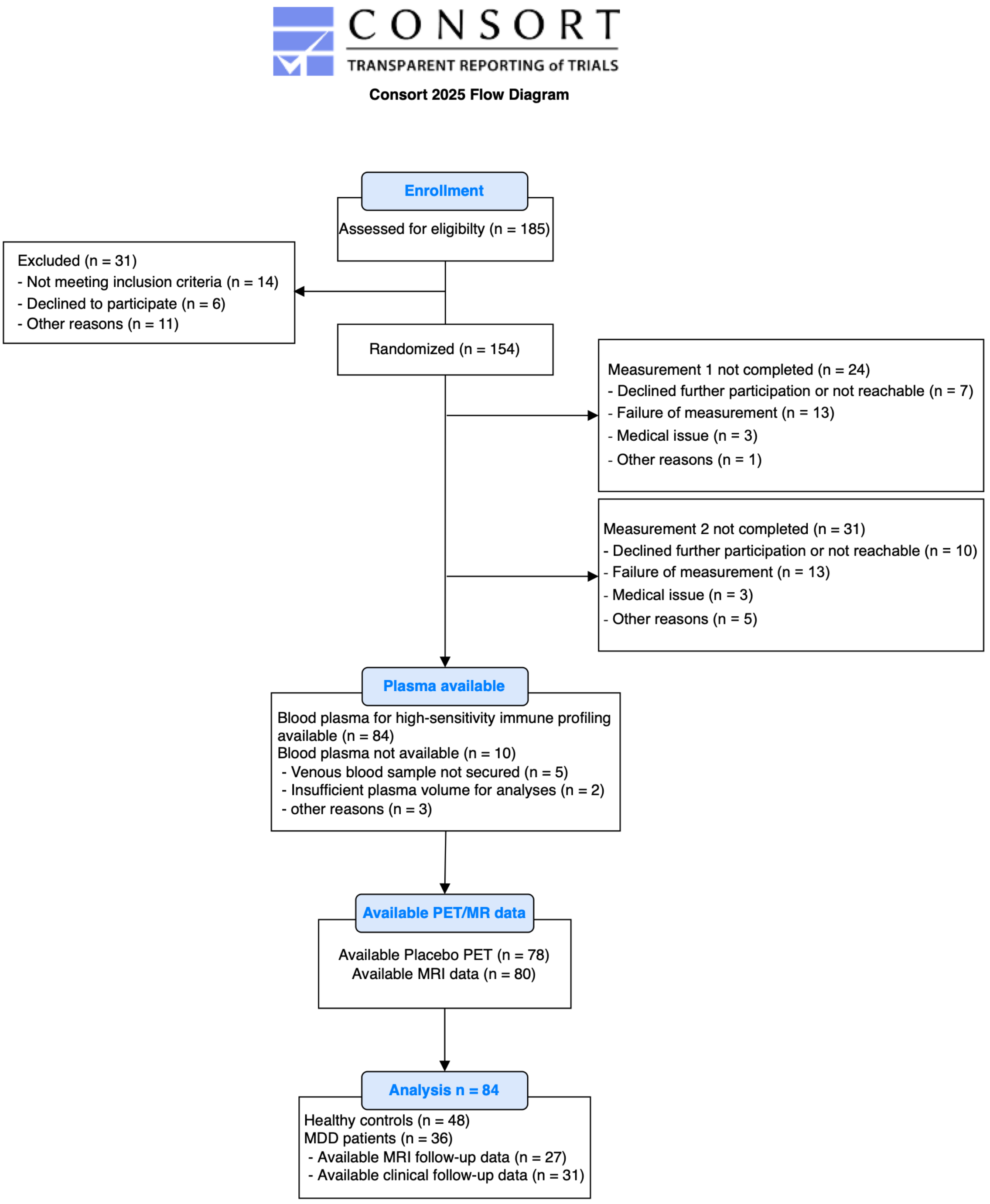

**Supplementary Figure S2: Replicate-level quality control of plasma immune marker measurements.** Concentrations of each analyte are shown on a log-scaled y-axis. Lines connect paired replicate measurements from the same sample. Points/lines are color-coded by replicate quality control (QC) status: black indicates samples passing the coefficient-of-variation criterion (gCV ≤20%), red indicates samples failing replicate precision (gCV >20%), orange indicates samples excluded because at least one replicate was below the limit of detection (LOD), and blue indicates singleton measurements or samples for which replicate precision could not be assessed. gCV, geometric coefficient of variation; GFAP, glial fibrillary acidic protein; IL, interleukin; LLOQ, lower limit of quantification; LOD, limit of detection; TNF, tumor necrosis factor.

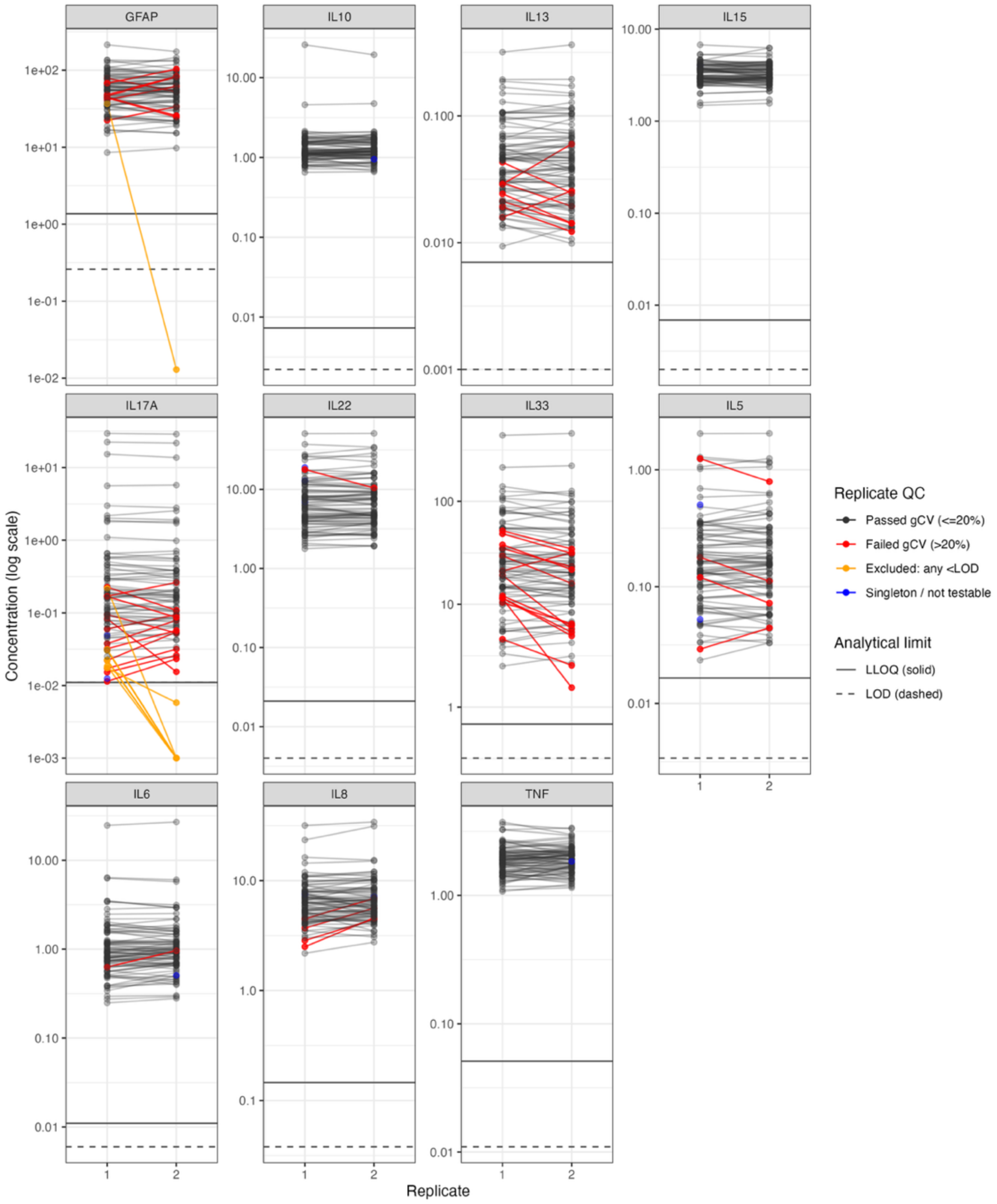

**Supplementary Figure S3: Plasma immune marker profile in major depressive disorder.** Group differences in immune marker concentrations. * = nominal $p < 0.05$, † = FDR-adjusted $p < 0.05$ within the domain. FDR, false discovery rate; GFAP, glial fibrillary acidic protein; IL, interleukin; TNF, tumor necrosis factor.

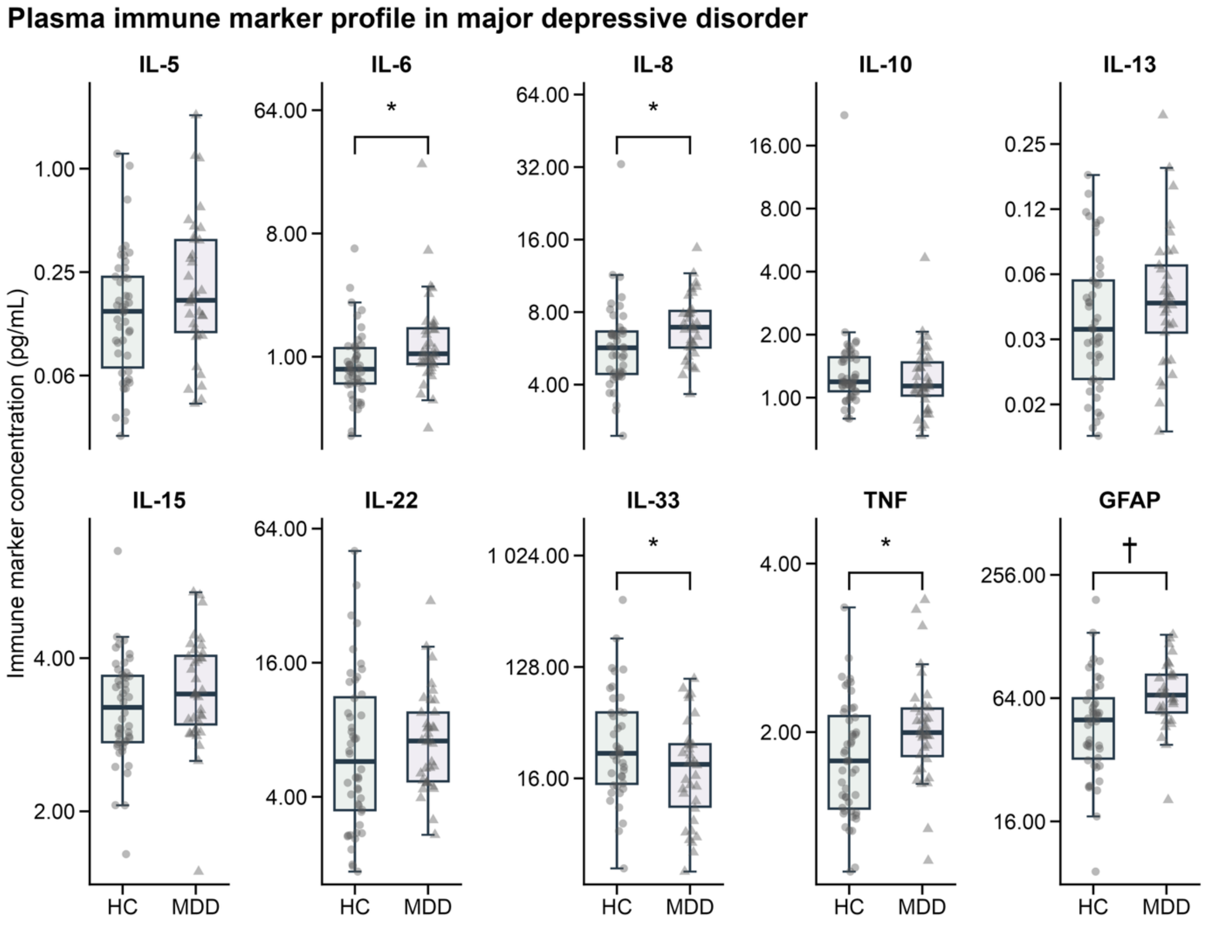

**Supplementary Figure S4: Diagnostic group-adjusted correlations between peripheral immune and routine laboratory markers.** Partial Spearman correlations were calculated while controlling for group. Only nominally significant correlations ($p < 0.05$, uncorrected for multiple comparisons) are displayed. CRP, C-reactive protein; GFAP, glial fibrillary acidic protein; IL, interleukin; TNF, tumor necrosis factor.

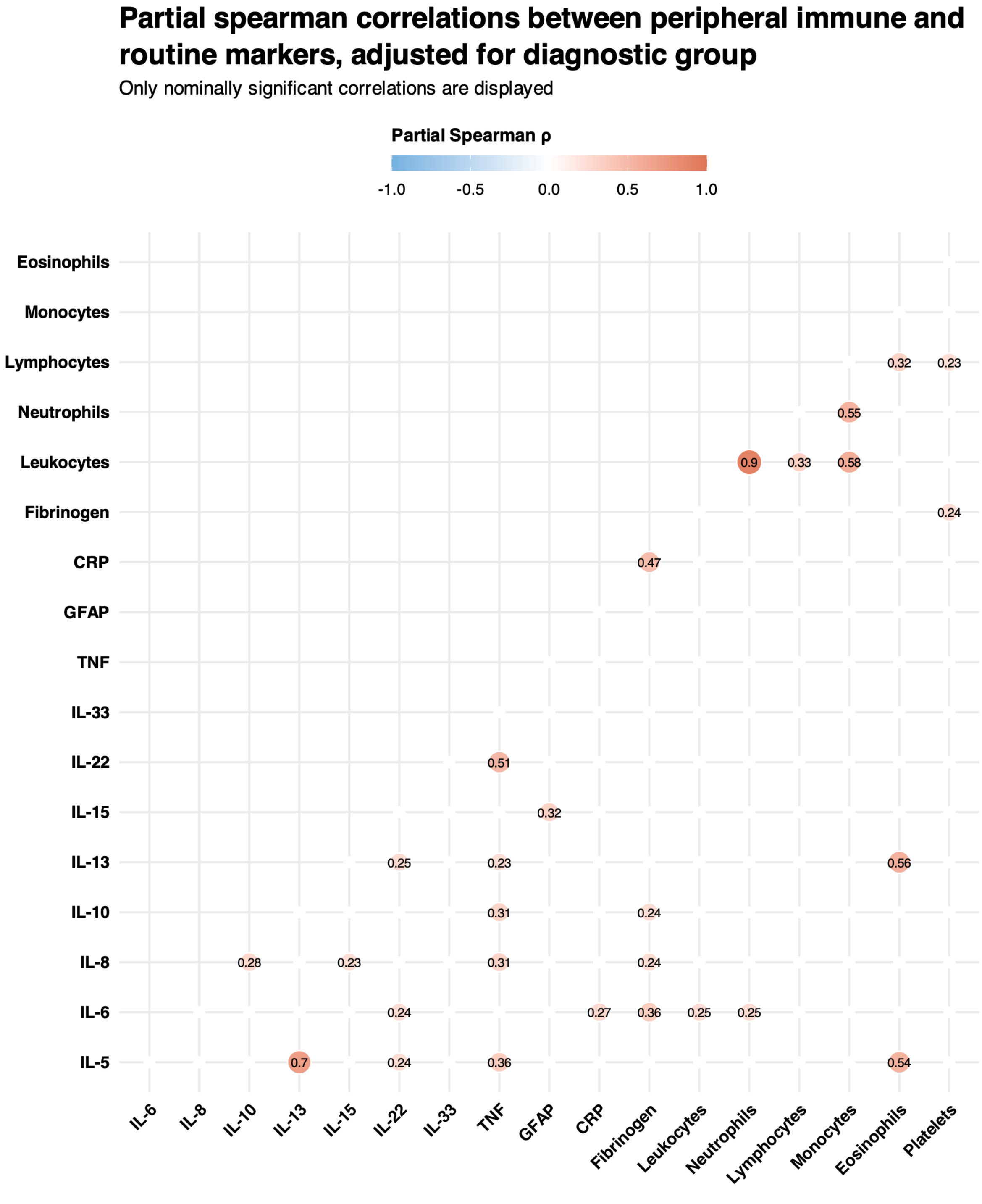

**Supplementary Figure S5: Association between symptom improvement and hippocampal volume change.** Greater six-week Montgomery-Åsberg Depression Rating Scale (MADRS) reduction was associated with more positive bilateral hippocampal-volume change among patients with complete longitudinal data ($n = 27$). * $p < 0.05$.

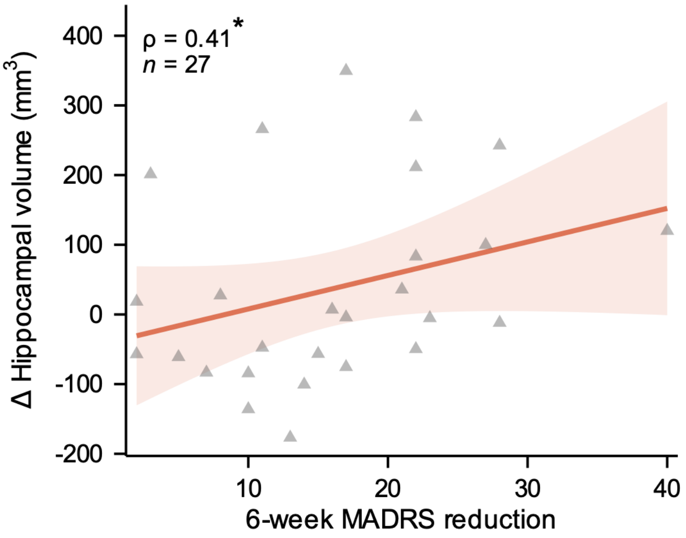
